# Effect of biannual azithromycin distribution on antibody responses to malaria, bacterial, and protozoan pathogens among children: A cluster-randomized, placebo-controlled trial in Niger

**DOI:** 10.1101/2021.04.23.21255957

**Authors:** Ahmed M. Arzika, Ramatou Maliki, E. Brook Goodhew, Eric Rogier, Jeffrey W. Priest, Elodie Lebas, Kieran S. O’Brien, Victoria Le, Catherine E. Oldenburg, Thuy Doan, Travis C. Porco, Jeremy D. Keenan, Thomas M. Lietman, Diana L. Martin, Benjamin F. Arnold, for the MORDOR-Niger Study Group

## Abstract

**Background:** The Macrolides Oraux pour Réduire les Décès avec un Oeil sur la Résistance (MORDOR) trial in Niger, Malawi, and Tanzania found that biannual mass distribution of azithromycin to children younger than 5 years led to a 13.5% reduction in all-cause mortality. Additional endpoints in the trial have attempted to elucidate the mechanisms for mortality reduction. In this pre-specified secondary analysis, we assessed the effect of azithromycin compared with placebo on IgG-based measures of infectious disease exposure with a multiplex bead assay that included antigens to malaria parasites (*Plasmodium falciparum, P. vivax, P. malariae, P. ovale*), bacterial pathogens (*Campylobacter* spp., enterotoxigenic *Escherichia coli*, *Vibrio cholerae*, *Salmonella enterica*, *Streptococcus pyogenes*) and protozoans (*Cryptosporidium parvum, Giardia duodenales*).

**Methods and Findings:** Thirty communities in rural Niger were randomized 1:1 to biannual distributions of azithromycin or placebo among children ages 1-59 months. The analysis included 5,642 blood specimens collected from 3,814 children ages 1-59 months, measured at 6, 12, 24, and 36 months of follow-up in a repeated cross-sectional design. *Campylobacter* spp. seroprevalence averaged over all study visits was lower in azithromycin communities compared to placebo (91% vs 94%, difference = –3%, 95% CI: –5%, –1%; *P*=0.03), which corresponded to a 29% lower seroconversion rate (1.30 versus 1.84 seroconversions per year, hazard ratio = 0.71, 95% CI: 0.56, 0.89; *P=*0.004). Antibody-based measures of infection with *P. falciparum* and group A streptococcus were consistently lower in azithromycin communities, but were not statistically different from placebo, and there were no other differences across pathogens. Strengths of the study included masking of participants, investigators, and analysts, high treatment coverage, large sample size, and objective outcomes. Principal limitations included the timing of blood collection with respect to treatment (approximately 6 months later, which could have missed transient effects in the weeks immediately following treatment), and the durability of IgG response following clearance of infection. Both limitations would lead the trial to under-estimate effects on antibody-based measures of infection.

**Conclusions:** The reduction in *Campylobacter* spp. despite these limitations suggests an effect on carriage, findings which align with an independent metagenomic analysis of rectal swabs collected in the same villages and with previously reported reductions in dysentery-associated mortality. Given significant sequelae of *Campylobacter* infection among preschool aged children, our results support at least one possible mechanism through which biannual mass distribution of azithromycin likely reduced mortality in this study population.

## Introduction

Child mortality rates in Niger are among the highest in Africa [1]. We previously conducted a cluster-randomized, placebo-controlled trial in Niger, Malawi, and Tanzania to assess the effect of biannual mass distribution of azithromycin on mortality among preschool aged children (MORDOR) [2]. In Niger, azithromycin reduced all-cause mortality by 18%, spurring interest in identifying specific mechanisms of pathogen reduction through which azithromycin reduced mortality. Follow-up analyses of MORDOR Niger demonstrated reductions in cause-specific mortality that spanned many leading causes of death in the region (e.g., malaria, pneumonia, diarrhea), suggesting effects were unlikely to result from a single mechanism [3]. An intensive monitoring trial in MORDOR Niger also documented lower levels of malaria parasitemia [4] and reduced *Campylobacter upsaliensis* carriage [5]. Studies that further clarify the mechanisms of mortality reduction from azithromycin could help identify complementary interventions or identify alternative interventions that lead to similar benefits but have lower potential to select for antimicrobial resistance [6].

In this pre-specified, secondary analysis of the MORDOR Niger trial, our objectives were to assess the effect biannual mass distribution of azithromycin to preschool aged children on serological measures of malaria, bacterial, and protozoan infections. Using a multiplex bead assay, we measured IgG responses to malaria parasites (*Plasmodium falciparum*, *P. vivax*, *P. malariae, P. ovale*), *Campylobacter* spp., enterotoxigenic *Escherichia coli* (ETEC), *Vibrio cholerae*, non-typhoidal *Salmonella* (serogroups B and D), *Streptococcus pyogenes* (serogroup A), *Cryptosporidium parvum*, and *Giardia duodenalis*. We hypothesized that, compared to placebo, children who received azithromycin would have lower levels of infection and thus lower IgG responses to the pathogens measured. Our rationale was based on evidence that azithromycin has antimicrobial activity against *P. falciparum*, *P. vivax*, group A *Streptococcus*, and gram negative bacteria including *Campylobacter*, ETEC, cholera, and *Salmonella* [7–14]. Although not a first line treatment, azithromycin has antimicrobial activity against enteric protozoans *Cryptosporidium* [15] and *Giardia* [16, 17].

## Materials and Methods

### Ethics Statement

The trial protocol was reviewed and approved by the Committee on Human Research at the University of California, San Francisco, and the Niger Ministry of Health’s Ethical Committee. Parents or guardians of enrolled children provided oral consent before each azithromycin or placebo treatment and at each specimen collection visit. Parents or guardians were instructed to report any adverse event within 7 days of treatment by contacting their village representative, who then reported events to the site coordinator and UCSF. An independent Data and Safety Monitoring Committee provided additional oversight. CDC researchers had access to de-identified samples for analysis (no personally identifying information).

### Study design and participants

MORDOR Niger was a cluster-randomized, placebo-controlled trial that randomized at the community level because of the intervention’s campaign-style, biannual mass distribution. Communities with 200 to 2,000 inhabitants based on the Niger 2012 census were eligible for inclusion in the trial, and children ages 1–59 months who weighed >3.8 kg were eligible for treatment. An intensive morbidity monitoring trial enrolled 30 communities and randomized them 1:1 to receive either biannual azithromycin or placebo to all children 1-59 months old (<u>NCT02048007</u>). The trial used a repeated cross-sectional design, whereby 40 children per community were randomly sampled in each measurement round and invited to participate in a monitoring visit. The trial’s open cohort design meant that children aged in and out of the study based on their age at the time of treatment. Field staff collected dried blood spots from participating children at baseline and annually thereafter at 12, 24, and 36 months of follow-up. The antibody substudy included a supplemental visit at 6 months, following the malaria season. Children who were randomly selected in multiple survey rounds contributed to longitudinal analyses.

### Randomization and Masking

Communities were randomized 1:1 using a sequence the trial biostatistician (TCP) generated. Unmasked members of the data team and Pfizer labeled the study drugs. Placebo and azithromycin had identical packaging to maintain masking. Participants, field staff, laboratory staff, analysts, and all investigators were masked to treatment assignments throughout the trial. Masked analyses were completed using a shuffled version of the treatment assignment variable [18]. Data were unmasked only after the final table and figure shells had been populated (documented through the article’s GitHub repository).

### Procedures

Children ages 1-59 months received azithromycin or identically-appearing placebo at the time of enrollment and every 6 months over the course of the study through community-wide census and MDA distributions performed by study staff. Children were given a volume of oral suspension equal to at least 20 mg per kilogram of body weight, which was measured by hanging scale for children unable to stand or by height stick for children who could stand, consistent with the Niger trachoma program. Children with known allergy to macrolides did not receive azithromycin or placebo.

### Antibody testing

#### Sample collection and preparation

Dried fingerprick blood spots (DBS) were collected onto calibrated filter paper wheels with 6 10µl extensions (TropBio Pty Ltd., Townsville, Queensland, Australia) and prepared as previously described [19]. Extensions were eluted overnight at 4°C in PBS containing 0.5% casein, 0.3% Tween-20, 0.5% polyvinyl alcohol, 0.8% polyvinylpyrrolidone, 0.02% sodium azide, and 3 µg/mL E. coli extract (Buffer B). Elutes were diluted to a final concentration of 1:400 with additional Buffer B to test on the multiplex bead assay.

#### Antigen coupling

Antigens were coupled to polystyrene beads (SeroMap Beads; Luminex Corporation, Austin, TX) through chemical modification as previously described [19]. Malaria antigens MSP-1, AMA1, Glurp-R0, LSA1, CSP, and HRP2 were coupled for the detection of *Plasmodium falciparum*. Species specific MSP-1 was used to detect *P. vivax*, *P. malariae, P. ovale* [20]. Bacterial and protozoan antigens from *Campylobacter jejuni* (p18, p39), enterotoxigenic *Escherichia coli* labile toxin B subunit (ETEC LTB), *Vibrio cholerae* toxin B subunit (CTB), *Salmonella* serogroups B and D (LPS), *Cryptosporidium parvum* (Cp17, Cp23), *Giardia duodenalis* (VSP-3, VSP-5), and *Streptococcus pyogenes* serogroup A Pyrogenic Exotoxin B (SPEB) were also coupled to beads using previously described methods [21, 22].

#### Multiplex bead assay

We measured IgG responses using a multiplex bead assay on the Luminex platform. Antigen-coupled beads (1250 per well/bead coupling) were incubated in 96-well assay plates with diluted sample for 1.5 hours then washed with 0.3% Tween-20 in PBS (PBST). Beads were washed with PBST and incubated with biotinylated mouse anti-human IgG and biotinylated mouse anti-human IgG4 for 45 minutes to detect antigen IgG bound to the beads. After additional washes with PBST, beads were incubated for 30 minutes phycoerythrin-labeled streptavidin to detect bound biotinylated anti-human IgG. After detection, beads were washed with PBST and incubated for 30 minutes with PBS containing 0.5% BSA, 0.05% Tween-20 and 0.02% sodium azide to remove loosely bound antibodies. After a final wash with PBST, beads were resuspended in PBS and stored at 4°C overnight. The next day, assay plates were read on a Bio-Plex 200 instrument (Bio-Rad, Hercules, CA) equipped with Bio-Plex manager 6.0 software (Bio-Rad). The median fluorescence intensity (MFI) with the background from the blank well (Buffer B alone) subtracted out (MFI-bg) was recorded for each antigen for each sample.

### Outcomes

We compared groups using geometric mean IgG responses, seroprevalence, and the seroconversion rate, including measurements at all follow-up times (6, 12, 24, 36 months). These were pre-specified, secondary outcomes for the trial (<u>NCT02048007</u>).

### Statistical analysis

#### Seropositivity cutoffs

We log_10_ transformed Luminex MFI-bg IgG levels before analysis. We converted IgG responses to seropositive and seronegative classes using seropositivity cutoffs derived from the mean plus 3 standard deviations (SD) of responses from a panel of known seronegative sera (malaria antigens), from ROC-derived cutoffs based on responses from known positive and negative specimens (*Cryptosporidium*, *Giardia*), or from the mean plus 3 SD of presumed unexposed measurements (all other antigens). We identified presumed unexposed measurements as those collected among children ≤12 months old that preceded a 10-fold increase in IgG in the longitudinal subsample [21]. For pathogens with multiple antigens measured, we classified children as seropositive if they were positive to any antigen. For *P. falciparum*, we examined individual antibody endpoints as well as a composite outcome, defined as a seropositive response to any *P. falciparum* antigen measured.

#### Age restrictions

We restricted age ranges included in the analyses based on pre-specified rules to exclude maternal IgG contributions and to focus the analysis on age ranges with heterogeneity in IgG responses. Before data were unmasked, we examined age-antibody profiles for each antigen and excluded from the primary analyses measurements among children <12 months (malaria responses) and <6 months (bacterial and protozoan responses) to remove potential maternally-derived IgG contributions (SI Figure 1, SI Figure 2) [23]. Additionally, we limited all analyses of ETEC LTB to ages 6-24 months and force of infection analyses based on seroconversion rates to measurements among children ≤24 months (all enterics except *Salmonella* sp.) because nearly all children older than 24 months were seropositive.

#### Estimation of mean differences

All comparisons were intention-to-treat. We compared mean differences between groups in geometric mean IgG levels and seroprevalence by pooling all post-treatment measurements. We estimated 95% confidence intervals using a non-parametric bootstrap that resampled communities with replacement (1000 iterations). We calculated exact permutation *P-*values from the randomization distribution of mean differences.

#### Age structured seroprevalence and force of infection

We used a current status, semi-parametric proportional hazards model to estimate force of infection from age-structured seroprevalence [24]. We fit a generalized additive mixed model with binomial errors and complementary log-log link:

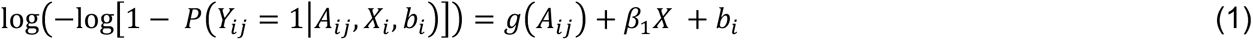

where*Y_ij_* is antibody seropositivity, *A_ij_* is age for child *j* in community *i*. *X_i_* is treatment allocation for community *i* (equal to 1 for azithromycin, 0 for placebo). The model included community-level random effects, *b_i_*, to allow for correlated outcomes within community. Function *g*(⋅) was parameterized with cubic splines that had smoothing parameters chosen through generalized cross-validation using the default in the R mgcv package [24]. The primary analysis pooled information over all post-randomization measurements available at the time of analysis (months 6, 12, 24, 36). We estimated the hazard ratio (HR) of seroconversion associated with biannual mass distribution of azithromycin as 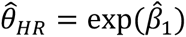.

We estimated age- and treatment-specific seroprevalence from the model as:

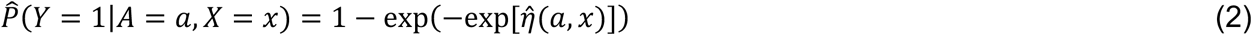

and we estimated age- and treatment-specific force of infection from the model as:

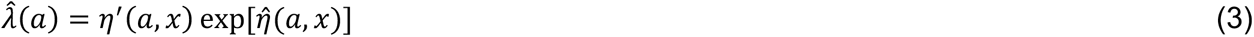

where *η*′(*a, x*) is the first derivative of the linear predictor from the complementary log-log model, *η*(*a, x*) [25]. We estimated *η*′(*a, x*) using finite differences from the model predictions [24, 26]. We estimated approximate, simultaneous 95% confidence intervals around age-specific seroprevalence and age-specific force of infection curves with a parametric bootstrap (10,000 replicates) from posterior estimates of the model parameter covariance matrix [27]. We used age-specific force of infection curves to visually confirm proportional hazards between groups. To estimate the marginal average force of infection in each group, we integrated over age [28].

#### Longitudinal rate estimates

A subset of children was sampled in multiple, repeated cross-sectional surveys and thus provided longitudinal antibody measurements (two to five visits). We conducted a supplemental analysis in this opportunistic subgroup to estimate prospective seroconversion and seroreversion rates. We defined seroconversion as an increase in IgG MFI-bg to a level above the antibody’s seropositivity cutoff. For pathogens with multiple measured antigens, a child was deemed to have seroconverted if either antibody response met the definition for seroconversion. We assumed that seroconversions occurred at the midpoint of the interval between measurements when estimating person-time at risk. We jointly estimated seroreversion rates using the same approach, using decrease in IgG across the seropositivity cutoff. We used a non-parametric bootstrap, resampling clusters with replacement, to estimate 95% confidence intervals for rate and incidence rate ratio estimates.

#### Subgroup analyses

We pre-specified examining treatment differences by age at the trial’s start date (<6 months vs older) and by trial phase (6, 12, 24, 36). We hypothesized that azithromycin could reduce antibody-based measures of transmission more among younger children who were immunologically naïve, and at later phases of the trial due to additional rounds of biannual MDA. We omitted a pre-specified subgroup analysis for rainy versus dry season because we ultimately determined that too few samples were collected after the rainy season (6-month samples only).

#### Statistical power, detectable effects, and adjustment for multiple comparisons

The MORDOR morbidity monitoring trial was designed around the primary antimicrobial resistance monitoring endpoints [5, 29]. For the present analyses, assuming a sample of 15 communities per arm and 140 measurements per community over four rounds, seroprevalence of 65% (*P. falciparum*), a community-level ICC of 0.004, and a two-sided alpha of 5%, we estimated that we would have 80% power to detect a reduction of 5.4 percentage points in seroprevalence due to intervention [30] (the pre-analysis plan provides additional details, https://osf.io/d9s4t/). Within each set of analyses, we estimated *P-*values adjusted for multiple comparisons allowing for a 5% false discovery rate using the Benjamini-Hochberg correction [31].

### Data and materials availability

Data and computational notebooks used to conduct the analyses are available through the Open Science Framework (https://osf.io/954bt) and Dryad (xx DOI forthcoming xx). Analyses used R statistical software, version 4.0.2.

## Results

### Study population and setting

The antibody substudy enrolled 3,814 children aged 1-59 months and tested a total of 5,642 blood specimens through the 36-month follow-up between March 2015 and June 2018 (Figure 1). One community withdrew from the trial at 36 months due to internal politics and trial fatigue. Except for the 6-month measurement, which took place after the seasonal rains, all other specimens were collected March – July toward the end of the dry season (SI Figure 3). Treatment coverage was high throughout the study, ranging from 71% to 93% of eligible children (Figure 1). At baseline, study arms were balanced across demographic characteristics and had similar seroprevalence to measured pathogens (Table 1). As reported in previous studies [21, 32], antibody responses to ETEC LTB and cholera CTB were highly cross-reactive (SI Figure 4). We excluded cholera CTB results from the primary analyses because we assumed that most of the responses reflected exposure to ETEC based on recent estimates of diarrhea etiology in nearby West African studies [33].

**Figure 1.**
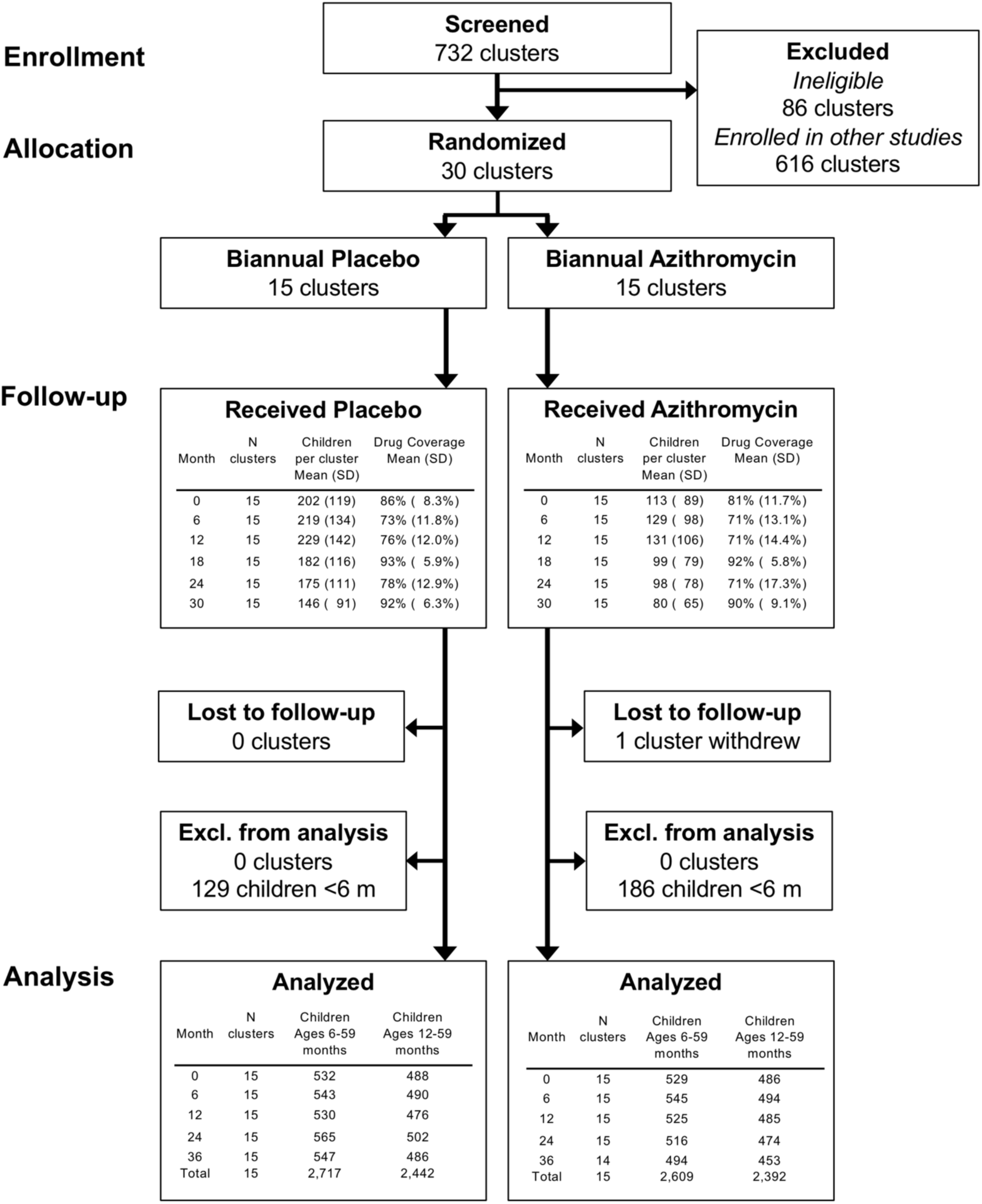
Study participant flow. 30 community clusters were randomly selected from a larger pool of clusters enrolled in the overall MORDOR Niger trial. A random sample of 40 children per community were selected for monitoring. Children <12 months old (malaria antigens) and <6 months (other antigens) were excluded from the primary analyses based on a pre-specified criteria to exclude maternal IgG contributions. Created with notebook https://osf.io/utxv3 .

**Table 1.** Baseline study group characteristics.

|  | Placebo<br>N = 15 | Azithromycin<br>N = 15 |
| --- | --- | --- |
| Number of children 0-59 months | 559 | 555 |
| Age, % |  |  |
| 0 y | 13 | 12 |
| 1 y | 14 | 14 |
| 2 y | 18 | 20 |
| 3 y | 26 | 24 |
| 4 y | 30 | 30 |
| Female, % | 45 | 48 |
| Malaria seroprevalence, % * |  |  |
| *P. falciparum* MSP-1<sub>19</sub> | 79 | 76 |
| *P. falciparum* AMA1 | 67 | 61 |
| *P. falciparum* GLURP-Ro | 26 | 21 |
| *P. falciparum* LSA1 | 11 | 8 |
| *P. falciparum* CSP | 5 | 4 |
| *P. falciparum* HRP2 | 2 | 2 |
| *P. malariae* MSP-1<sub>19</sub> | 13 | 12 |
| *P. ovale* MSP-1<sub>19</sub> | 2 | 3 |
| *P. vivax* MSP-1<sub>19</sub> | 1 | 2 |
| Bacteria and protozoa seroprevalence, % † |  |  |
| *Campylobacter* sp. p18 or p39 | 92 | 91 |
| ETEC LTB | 88 | 84 |
| *Salmonella* sp. LPS serogroups B or D | 48 | 45 |
| *Cryptosporidium* sp. Cp17 or Cp23 | 85 | 83 |
| *Giardia* sp. VSP-3 or VSP-5 | 77 | 76 |
| *Streptococcus* sp. serogroup A SPEB | 63 | 60 |

\* Malaria antibody seroprevalence estimated among children ages 12-59 months (baseline Placebo *N* = 488, Azithromycin *N* = 486), per the age group used in the primary analysis.
† Bacteria and protozoan antibody seroprevalence estimated among children ages 6-24 months (ETEC LTB, baseline Placebo *N* = 201, Azithromycin *N* = 202) or among children ages 6-59 months (all others, baseline Placebo *N* = 532, Azithromycin *N* = 539) or, per the age groups used in the primary analysis.
‡ Created with notebook https://osf.io/qeg4u

### Effects on malaria antibody response

Children had high IgG seroprevalence to *P. falciparum* MSP-1_19_ and AMA1 antigens, evidence of limited exposure to *P. malariae*, and evidence of very low exposure to *P. vivax* or *P. ovale* (Figure 2A). There was heterogeneity between study communities in the longer-lived MSP-1_19_ and AMA1 antibodies, and overall seroprevalence to shorter-lived *P. falciparum* antibodies (GLURP-Ro, LSA1, CSP, HRP2) was considerably lower. IgG responses by age exhibited a characteristic pattern of waning up to 12 months, due to loss of maternal antibodies, with monotonic increases thereafter (SI Figure 1). Children who received azithromycin had a transient reduction in *P. falciparum* IgG seroprevalence between ages 12-36 months (Figure 2B), but the overall difference between groups from ages 12-59 months was small (–4% difference, 95% CI:–12%, +3%; *P=*0.32). This corresponded to a 12% relative reduction in the hazard of seroconversion to any of the *P. falciparum* antigens (HR = 0.88, 95% CI: 0.62, 1.26). Antigen-specific differences between groups showed small, non-statistically significant reductions among children who received azithromycin based on seroprevalence (Figure 2C) and force of infection measured by the seroconversion rate (Figure 2D). Antigen-specific, age-seroprevalence curves showed similar overall patterns between groups, with largest reductions in *P. falciparum* AMA1 (SI Figure 5), consistent with comparisons of community-level means (SI Figure 6).

**Figure 2.**
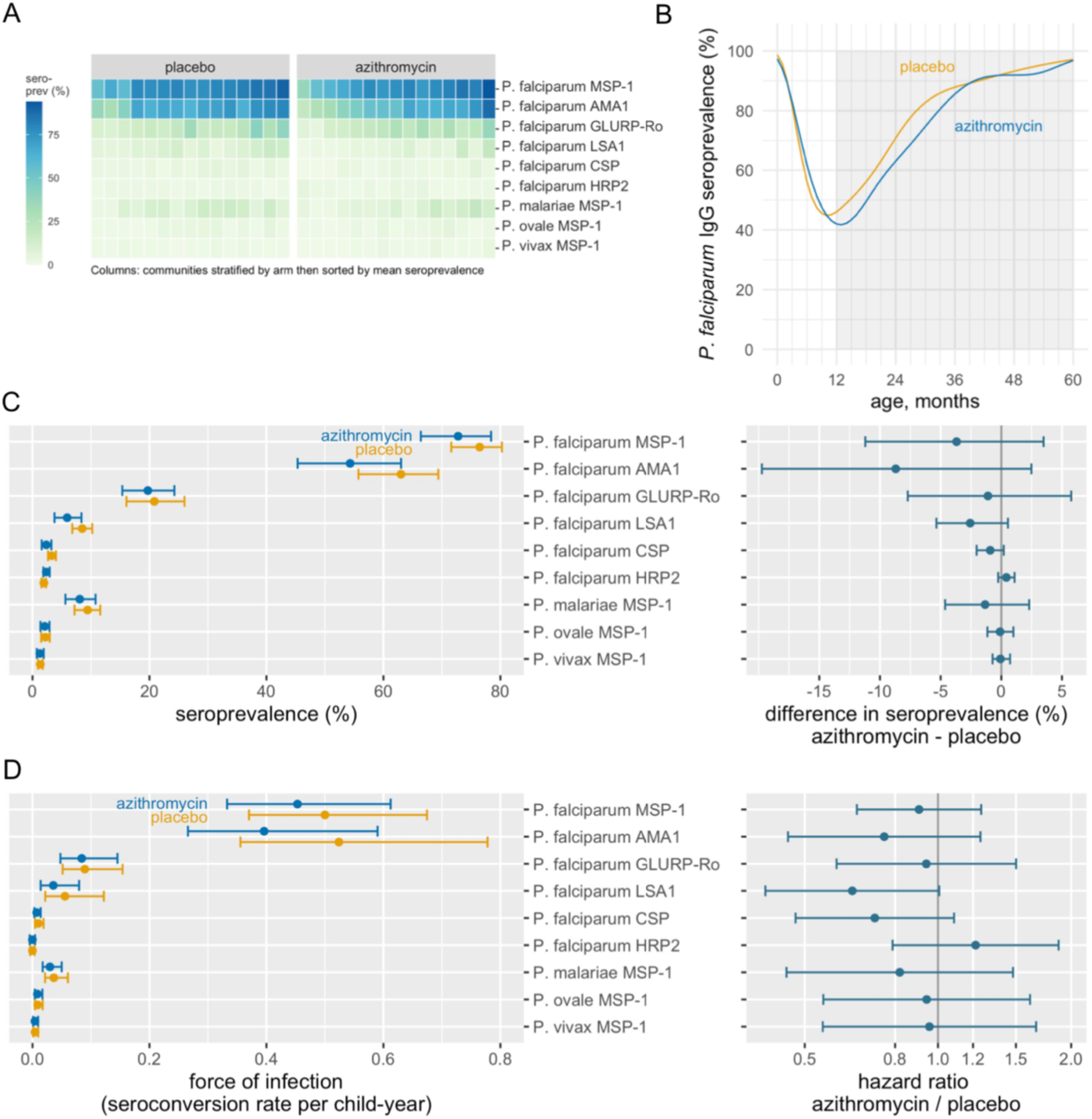
Malarial IgG antibody responses among preschool aged children in the MORDOR Niger trial, 2015-2018. **(A)** Community-level seroprevalence to malarial antigens. Columns represent individual communities, which were stratified by treatment group and then sorted by overall mean seroprevalence. **(B)** Seroprevalence to *P. falciparum* (positive to any measured antigen) by age and treatment arm, estimated with semiparametric splines. The shaded region from 12-59 months indicates the age range included in the primary analysis. **(C)** Antigen-specific IgG seroprevalence by treatment arm and difference between arms. **(D)**. Antigen-specific force of infection estimated by the seroconversion rate, and hazard ratio for comparison between arms. No between-group comparisons were statistically significant at the 95% confidence level after FDR correction. Created with notebooks https://osf.io/b2v3r, https://osf.io/37ybm, https://osf.io/fwxn5, which include detailed point estimates, and additional, consistent results based on geometric mean IgG levels.

### Effects on bacterial and protozoan antibody response

*Campylobacter* and ETEC seroprevalence were >90% among children 6-24 months, with little heterogeneity between communities (Figure 3A). *Salmonella* serogroups B and D, *Streptococcus* serogroup A, *Cryptosporidium*, and *Giardia* seroprevalence was lower compared to the highest transmission pathogens and more heterogeneous between communities (Figure 3A). Many bacterial and protozoan pathogens showed evidence of maternal IgG contributions through age six months, and mean *Campylobacter* and *Giardia* IgG levels declined modestly beyond ages 18-24 months (SI Figure 2).

**Figure 3.**
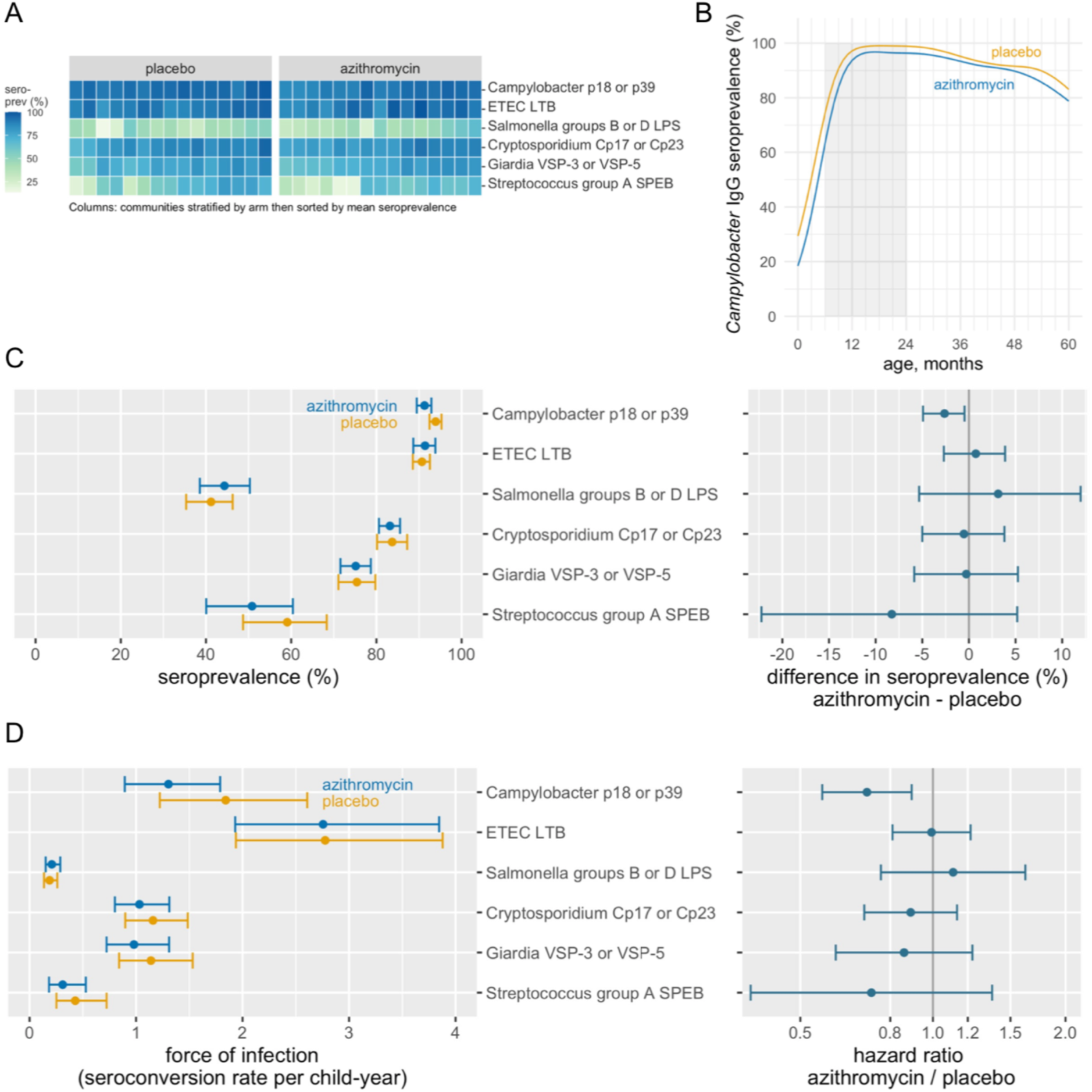
Bacterial and protozoan IgG antibody responses among preschool aged children in the MORDOR Niger trial, 2015-2018. **(A)** Community-level seroprevalence to malarial antigens. Columns represent individual communities, which were stratified by treatment group and then sorted by overall mean seroprevalence. **(B)** Seroprevalence to *Campylobacter* spp. p18 or p39 antigens by age and treatment arm, estimated with semiparametric splines. The shaded region from 6-24 months indicates the age range included in the force of infection analyses, based on a pre-specified rule. **(C)** Pathogen-specific IgG seroprevalence by treatment arm and difference between arms. **(D)**. Pathogen-specific force of infection estimated by the seroconversion rate, and hazard ratio for comparison between arms. The reduction in *Campylobacter* spp. transmission remained statistically significant after FDR correction. Created with notebooks https://osf.io/b2v3r, https://osf.io/smwbn, https://osf.io/fwxn5, which include detailed point estimates and additional, consistent results based on geometric mean IgG levels.

*Campylobacter* seroprevalence was lower among children who received azithromycin compared with placebo (91% vs 94%, difference = –3%, 95% CI: –5%, –1%; *P*=0.03) (Figure 3C), and age-seroprevalence curves showed a consistent reduction across all ages (Figure 3B). We estimated 0.5 fewer *Campylobacter* seroconversions per child-year at risk, a 29% relative reduction (1.30 versus 1.84 seroconversions per year), based on a semiparametric proportional hazards model (HR = 0.71, 95% CI: 0.66, 0.89; *P=*0.004), which remained significant after correction for multiple testing (Figure 3D). There were no statistically significant reductions in seroprevalence or force of infection for other measured bacterial and protozoan pathogens. Age-seroprevalence curves (SI Figure 7) and community level means (SI Figure 8) were consistent with the overall results.

### Additional analyses

Children included in multiple cross-sectional samples between ages 12-59 months contributed to longitudinal analyses of malaria seroconversion (919 children, 2,197 measurements). Longitudinal samples from children ages 6-59 months (1,038 children, 2,516 measurements) contributed to analyses of *Salmonella* and *Streptococcus*, and longitudinal samples among children 6-24 months (313 children, 680 measurements) contributed to analyses of the remaining enteric pathogens. Seroconversion rates were generally higher when estimated longitudinally compared with those estimated from age-structured seroprevalence. Longitudinal analyses demonstrated larger reductions in *P. falciparum* seroconversion rates among children who received azithromycin compared to the primary analysis (e.g., AMA1 seroconversion incidence rate ratio: 0.49, 95% CI: 0.31, 0.76), but most comparisons were slightly underpowered given the small size of the longitudinal cohort (SI Table 1). Longitudinal comparisons of bacterial and protozoan pathogens were largely consistent with the primary analysis but were also slightly underpowered. For example, the *Campylobacter* seroconversion rate was 19% lower (IRR: 0.81, 95% CI: 0.62, 1.04) in the azithromycin group compared to placebo in the longitudinal analysis (SI Table 1). Longitudinal analyses showed seroreversion across all antibodies measured except for responses to *Campylobacter* and ETEC, with some evidence for higher *Cryptosporidium* and *Giardia* seroreversion rates in the azithromycin group (SI Table 2).

There was no evidence for effect modification by study phase (SI Figure 9) or child age at trial start (SI Figure 10).

## Discussion

In this analysis of IgG antibody response to malarial, bacterial, and protozoan pathogens we found that biannual mass distribution of azithromycin to children 1-59 months reduced *Campylobacter* seroprevalence and seroconversion rates. Overall, azithromycin had limited effects on antibody-based measures of pathogen exposure against a backdrop of very high transmission for most pathogens. Antibody responses showed that infection was extremely common for most pathogens studied, for example, seroprevalence to *Campylobacter* and ETEC was close to 100% by age 18 months, and was >90% for *P. falciparum* by age 40 months.

A reduction in IgG responses to *Campylobacter jejuni* among children who received azithromycin is consistent with the trial’s earlier report of reduced mortality in azithromycin-treated communities attributed to dysentery [3]. The results are also consistent with a separate metagenomic deep sequencing analysis of rectal swabs collected from children in the same communities, which showed a reduction in *Campylobacter upsaliensis* carriage but no other differences in gut microbiome composition among children in azithromycin-treated communities [5]. IgG responses p18 and p39 could reflect previous infections from multiple *Campylobacter* species. A rabbit polyclonal antibody raised against recombinant p18 antigen cross reacted with an 18-kDa protein in cell extracts from a broad selection of other *Campylobacter* species [34], and the high level of amino acid sequence identity predicted by a Basic Local Alignment Search Tool for Proteins (BLASTP) analysis [35, 36] of the *C. jejuni* and *C. upsaliensis* p18 and p39 antigens (88% and 80%, respectively) suggests that measured IgG responses could reflect previous infection by one or both species (SI Table 3). Although we estimated that azithromycin treatment reduced the hazard of *Campylobacter* seroconversion by 29%, by age 18 months more than 95% of children in both groups were seropositive to *Campylobacter*, reflecting very high force of infection. These results are consistent with eight, high-resolution, longitudinal cohorts that found 85% of children in low-resource settings had experienced at least one *Campylobacter* infection by age 12 months [37]. *Campylobacter* infections among preschool aged children have significant clinical sequelae including acute bloody diarrhea (identified as the leading cause) [38], increases in intestinal inflammatory markers [37], and subsequent growth failure [39]. As we describe below, features of this study could have led us to under-estimate effects of the intervention on pathogen carriage, so effects demonstrated on *Campylobacter* force of infection despite built-in conservatism in the estimates suggest a potentially important mechanism for the intervention’s effect on mortality in Niger.

This study illustrates how multiplex antibody assays can measure intervention effects across diverse pathogens at relatively low cost and from samples that are relatively easy to collect [40]. Multiplex assays are especially valuable for interventions like azithromycin, which could plausibly have broad-based impacts. Previous results from this trial found that azithromycin reduced mortality attributed to malaria, diarrhea, dysentery, and pneumonia [3], and reduced malaria parasitemia [4]. Yet, our finding of no clear reduction in IgG responses to malaria and bacterial pathogens beyond *Campylobacter* suggests that in high transmission settings, infrequent IgG measurements by themselves are potentially an insensitive trial endpoint. Azithromycin has modest activity against *P. falciparum* through action against the parasite’s apicoplast [7, 8], and high efficacy against *P. vivax* both as a prophylactic to prevent infection [9] and as a treatment therapy [10]. Azithromycin has broad-spectrum activity against gram-positive and atypical bacteria and has been shown to be highly effective for treatment of bacterial enteric pathogen infections, including enterotoxigenic *Escherichia coli* (ETEC), *Vibrio cholerae*, and non-typhoidal *Salmonella enterica* [12–14]. Several trials have demonstrated that azithromycin has comparable efficacy to penicillin in treating group A streptococcal pharyngitis [11]. Azithromycin has been shown to have modest anti-parasitic activity against *Cryptosporidium parvum* [15]. Less is known about its effect on *Giardia duodenalis* infections in humans, but *in vitro* and *in vivo* animal studies have shown azithromycin had comparable antimicrobial activity against *G. duodenalis* to metronizadole (the first line treatment) [16] and small scale (uncontrolled) human studies suggest cure rates in excess of 90% [17]. Yet, even if azithromycin successfully treated infections from these pathogens, IgG responses might be an insensitive marker of efficacy if treatment did not reduce the immune response or if a child was reinfected between the 6-month community-based treatments, leading to a boost in IgG levels. Additionally, IgG longevity or low-density infections that elicited a robust adaptive immune response could explain the small reductions observed in *P. falciparum* IgG responses despite lower levels of parasitemia and parasite density in the azithromycin group [4].

This study had limitations, all of which could have led us to under-estimate effects of azithromycin on pathogen-induced IgG response. Most samples were collected at the end of the dry season, many months after peak malaria season, which coincides with seasonal rains (SI Figure 3). Subgroup analyses by study phase compared groups at 6-months after enrollment, immediately following the rainy season, and showed no consistently larger effect across antigens at that timepoint (SI Figure 9), suggesting that measurement timing alone would not explain the absence of effects on malaria and other pathogens. A second limitation was that the study collected blood spots approximately 6 months after each treatment administration. If azithromycin had short term effects on infection and antibody response, without effects on longer term pathogen carriage, then this study could have missed such transient effects. The timing of monitoring visits was chosen to provide longer-term data about community antibiotic resistance (i.e., resistance that persisted even months after the mass drug administration) [6]. However, azithromycin would be expected to have its strongest effect within days or weeks of administration. An analysis of timing of mortality in MORDOR Niger suggested that most of the intervention effect occurred within three months of distribution [41]. Among children ages 0-24 months, macrolides have been shown to reduce the risk of *Campylobacter* detection in stool significantly 0-15 days after treatment with reduced protection through 45 days, and no protection thereafter [37]. A third limitation was that the primary analysis followed the repeated cross-sectional design and thus did not account for high levels of between-child variability in IgG response. Repeated cross-sectional samples enabled the trial to continually enroll young children as the study progressed and was protected from potential bias through loss-to follow-up. Nevertheless, longitudinal analyses on a subset of the children who provided multiple specimens over time had similar precision as the primary analysis, illustrating much higher efficiency of longitudinal designs for IgG responses. Despite this limitation, high levels of agreement between the repeated cross-sectional and longitudinal analyses provided a valuable internal consistency check. Finally, the campaign-style distribution of azithromycin every 6 months meant that some children would have been up to 6-months old before their first treatment. For very high transmission pathogens, it is likely that many children had already been infected and may have had a robust IgG response that was insensitive to reduced pathogen carriage following azithromycin treatment.

In conclusion, this study demonstrated that biannual mass distribution of azithromycin reduced antibody-based measures of *Campylobacter* infection, consistent with independent metagenomic analyses in the same study communities. There was no evidence for significant reductions in IgG antibody-based measures of infection with malaria or other measured bacterial and protozoan pathogens. Given the clinically significant sequelae from *Campylobacter* infection among preschool-aged children, our results point to at least one important mechanism through which azithromycin plausibly reduced mortality in Niger. Studies of infection in the weeks immediately following treatment may provide additional insights into the mechanisms through which mass distribution of azithromycin reduces child mortality.

## Acknowledgements

### Funding

This work was supported by the Bill & Melinda Gates Foundation (award no. OPP1032340 to TML) and was supported in part by an unrestricted grant from Research to Prevent Blindness and by the National Institute of Allergy and Infectious Diseases (award no. K01-AI119180 to BFA). The Gates Foundation approved the study design, but had no role in data collection, data analysis, data interpretation, or writing of the report.

### Author contributions

following CRediT taxonomy, conceptualization (DLM, ER, TML, JDK, BFA), data curation (JDK, VL, BFA), formal analysis (BFA), funding acquisition (TML), investigation (DLM, ER, JWP, BFA), methodology (BFA), project administration (AMA, RM, EL, TML), resources (JWP, DLM, ER, EBG), software (BFA, TCP, VL), supervision (AMA, RM, EL, KSO), validation (EBG, ER), visualization (BFA), Writing – Original Draft Preparation (AMA, BFA, DLM, EBG), Writing – Review & Editing (AMA, RM, EBG, ER, JWP, EL, KSO, VL, CEO, JDK, TCP, TD, TML, DLM, BFA).

## MORDOR-Niger Study Group Investigators

***University of California, San Francisco, San Francisco, CA, USA***

Benjamin F Arnold, Catherine Cook, Sun Y Cotter, Thuy Doan, Dionna M Fry, Jeremy D Keenan, Victoria Le, Elodie Lebas, Thomas M Lietman, Ying Lin, Kieran S O’Brien, Catherine E Oldenburg, Travis C Porco, Kathryn J Ray, Philip J Rosenthal, George W Rutherford, Benjamin Vanderschelden, Nicole E Varnado, Lina Zhong, Zhaoxia Zhou

***The Carter Center, Atlanta, GA, USA***

E Kelly Callahan, Aisha E Stewart

***The Carter Center Niger, Niamey, Niger***

Ahmed M Arzika, Sanoussi Elh Adamou, Nana Fatima Galo, Fatima Ibrahim, Salissou Kane, Mariama Kiemago, Ramatou Maliki

***Programme National de Santé Oculaire, Niamey, Niger***

Amza Abdou, Boubacar Kadri, Nassirou Beido

***Johns Hopkins University, Baltimore, MD, USA***

Jerusha Weaver, Sheila K West

***International Trachoma Initiative, Decatur, GA, USA***

Paul M Emerson

**Steering Committee.** The steering committee for the trial consisted of Robin L Bailey, Jeremy D Keenan, Thomas M Lietman, Travis C Porco, and Sheila K West.

**Sponsor program officers.** The program officers from the trial’s sponsor included Rasa Izadnegahdar, Julie Jacobson, Thomas Kanyok, and Erin Shutes (Bill & Melinda Gates Foundation, Seattle, WA, USA).

**Data and Safety Monitoring Committee (DSMC).** The trial’s DSMC included Judd L Walson (University of Washington, Seattle, WA, USA), Allen W Hightower (Centers for Disease Control and Prevention, Atlanta, GA, USA), Emily E Anderson (Loyola University, Chicago, IL, USA), Wondu Alemayehu (Fred Hollows Foundation, Addis Ababa, Ethiopia), and Latha Rajan (Tulane University, New Orleans, LA, USA).

## Supplementary Information Materials

**SI Figure 1.**
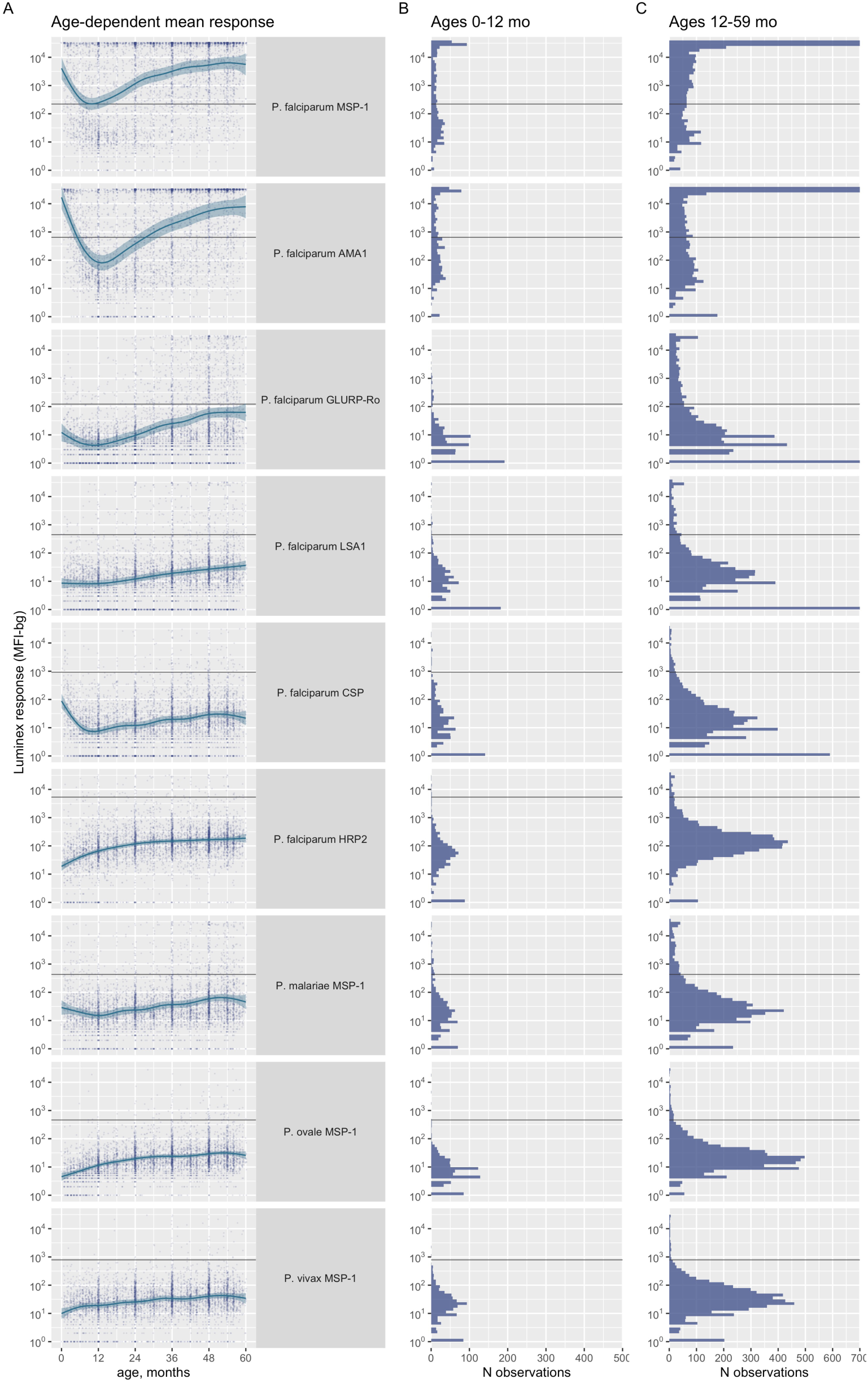
Malarial antibody responses among children ages 0-59 months in MORDOR Niger. (**A**) Age dependent mean response estimated with cubic splines, shaded bands are simultaneous 95% confidence intervals. Distribution of antibody response among children ages 0-12 months (**B**), and ages 12.1 to 59 months (**C**). Horizontal lines mark the seropositivity cutoffs derived from panels of negative sera. Created with notebook https://osf.io/vqtbm.

**SI Figure 2.**
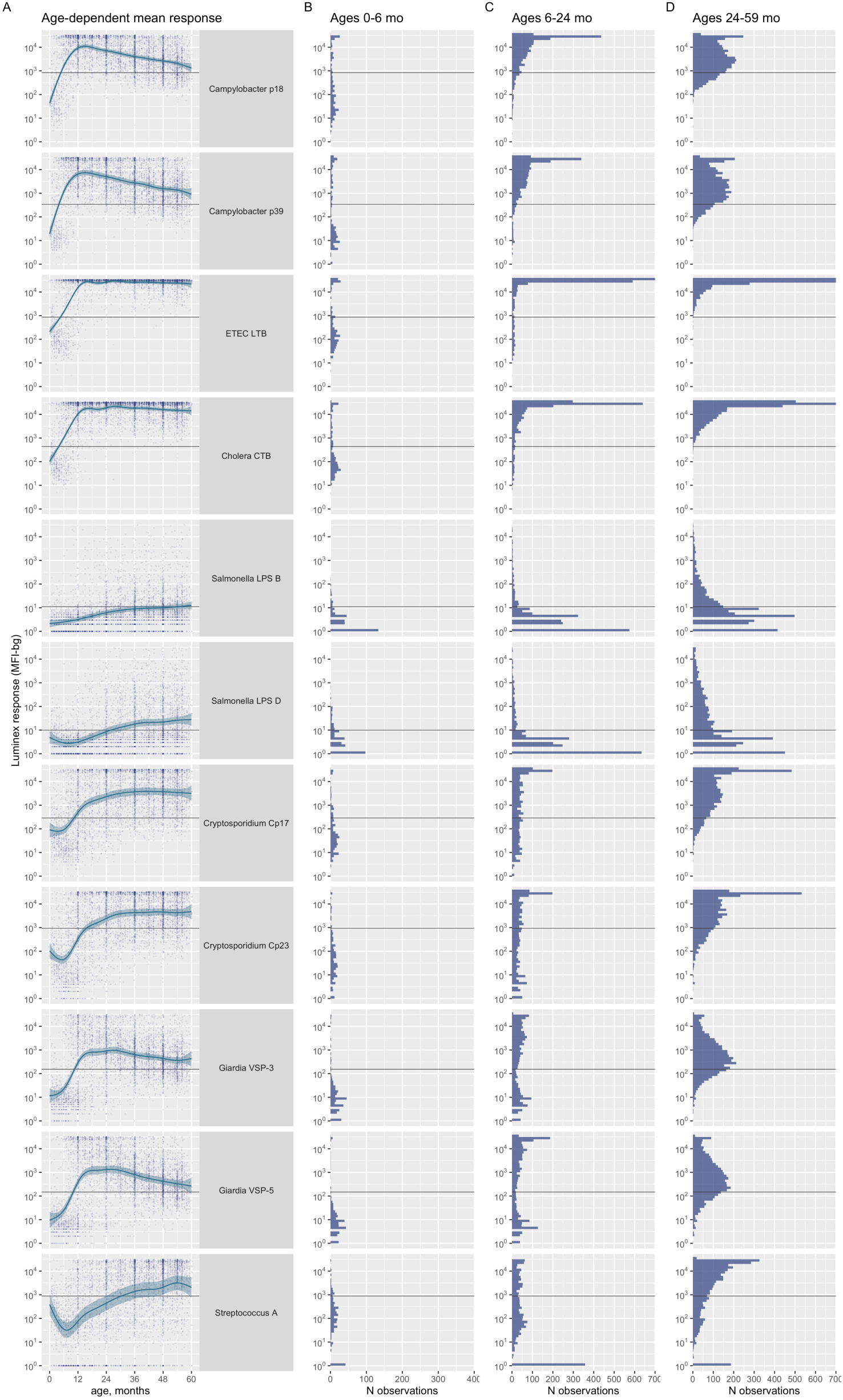
Antibody responses to bacteria and protozoan pathogens among children ages 0-59 months in MORDOR Niger. (**A**) Age dependent mean response estimated with cubic splines, shaded bands are simultaneous 95% confidence intervals. Distribution of antibody response among children ages 0-6 months (**B),** ages 6.1 to 24 months (**C**), and 24.1 to 59 months (**D**). Horizontal lines mark the seropositivity cutoffs derived through ROC curves (*Cryptosporidium* sp., *Giardia* sp.) or using the mean plus 3xSD among presumed unexposed children (others, described in Methods). Created with notebook https://osf.io/vqtbm.

**SI Figure 3.**
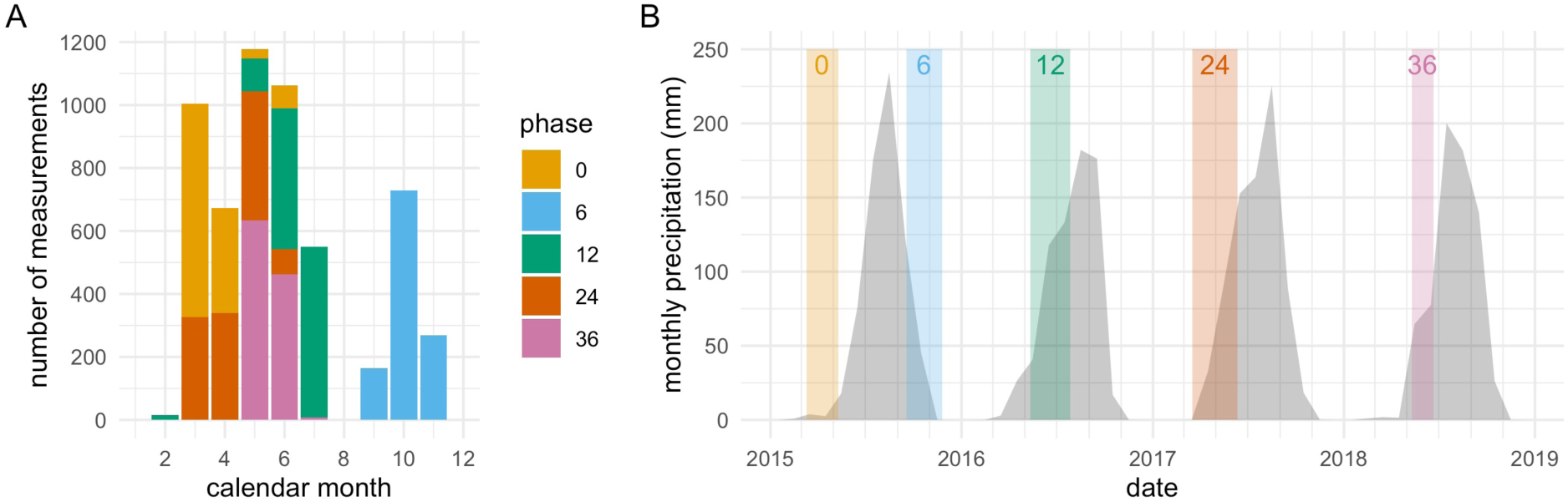
Dried blood spot measurement timing and monthly rainfall in the MORDOR Niger trial. **(A)** Specimens collected by calendar month and study phase (months since baseline). **(B)** Monthly precipitation in study communities. Colored shading indicates timing of dried blood spot specimen collection for each study phase. Created with notebook https://osf.io/pem3z .

**SI Figure 4.**
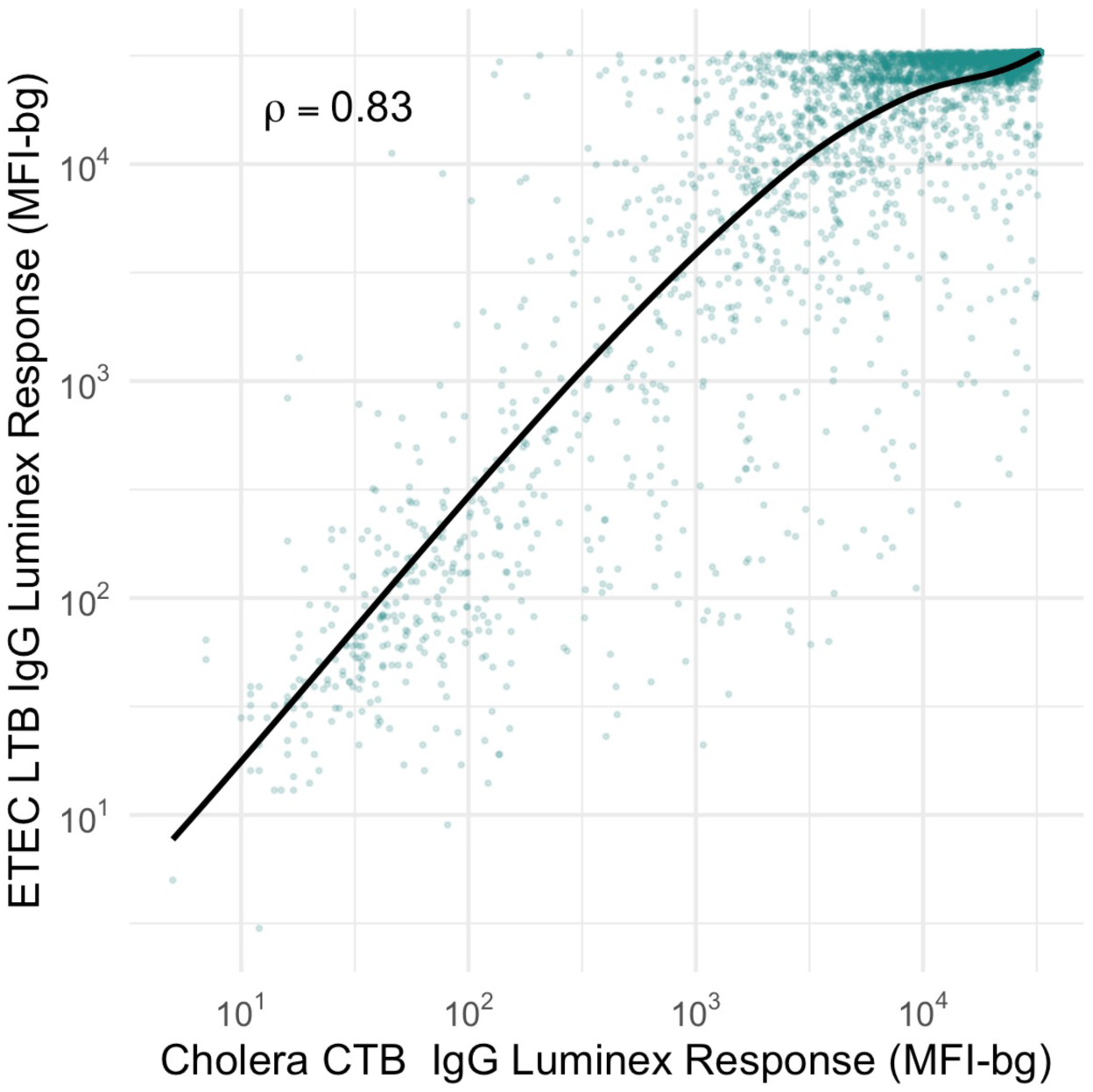
Correlation between antibody responses to enterotoxigenic *E. coli* labile toxin B subunit (ETEC LTB) and *V. cholera* toxin B subunit (cholera CTB) among children ages 6-59 months in Niger. Spearman rank correlation (!) estimated from 5,642 measurements (points). Solid line is a locally weighted smooth. Created with notebook https://osf.io/5vs7r.

**SI Figure 5.**
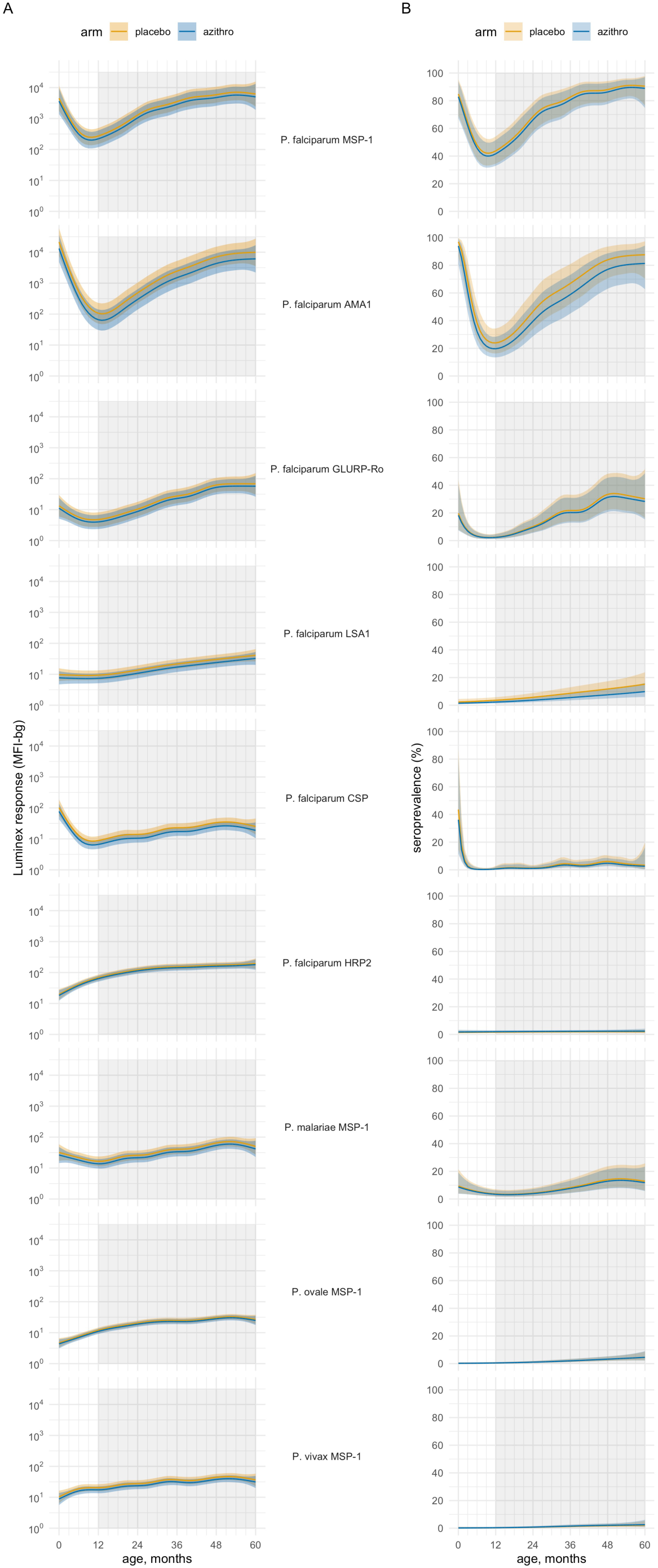
Malarial IgG antibody response among preschool aged children in the MORDOR Niger trial by treatment group and age, 2015-2018. **(A)** Mean IgG responses (MFI-bg) **(B)** Seroprevalence. Shaded area indicates age range included in analyses (12-59 months). Group means estimated using semiparametric cubic splines. Shaded bands are simultaneous 95% confidence intervals. Created with notebook https://osf.io/smwbn .

**SI Figure 6.**
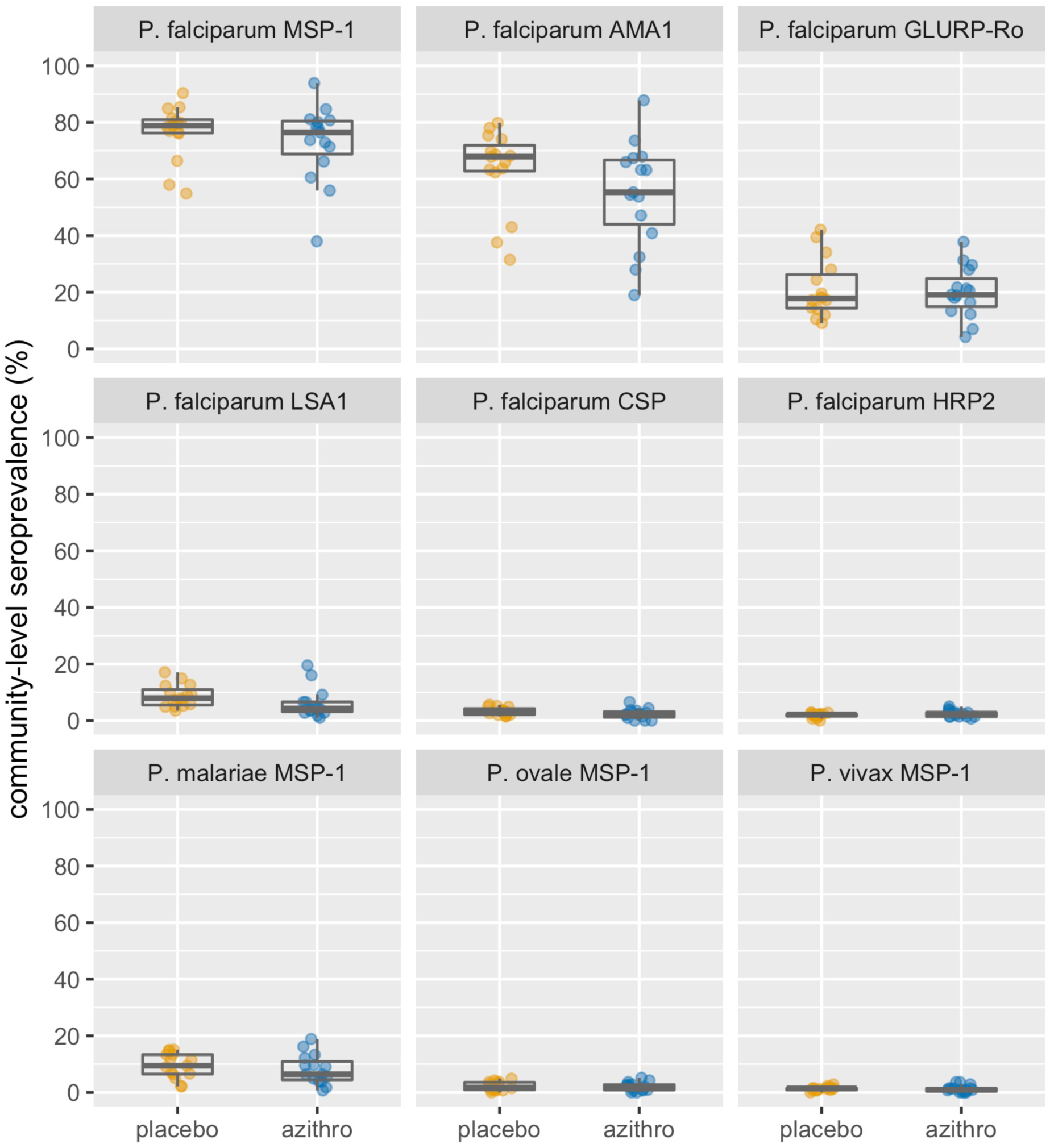
Community level IgG seroprevalence for malarial antigens among preschool aged children in the MORDOR Niger trial by treatment group, 2015-2018. Box plots show the median and interquartile range for the 15 communities in each group. Created with notebook https://osf.io/b2v3r .

**SI Figure 7.**
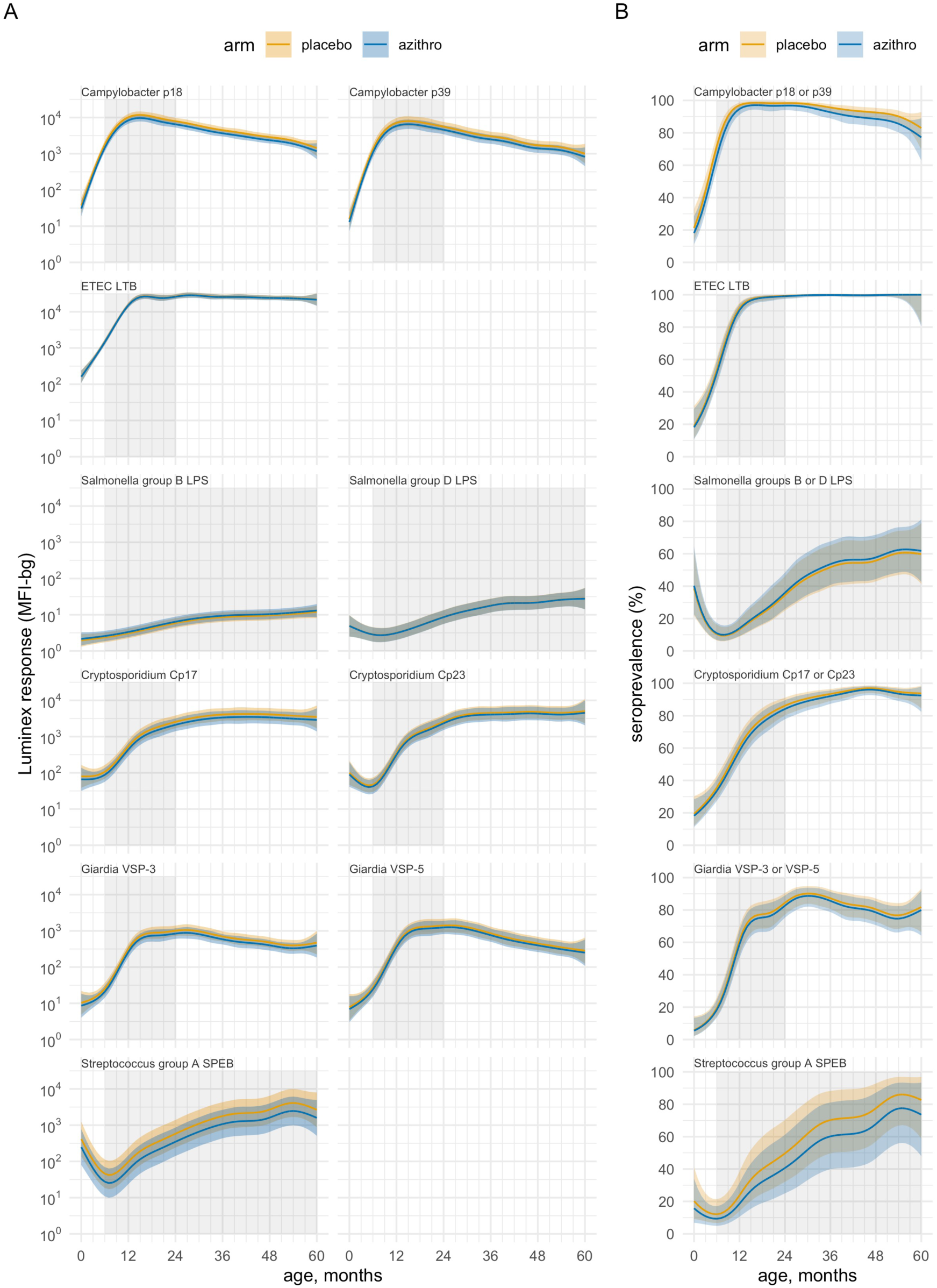
IgG antibody responses to bacterial and protozoan pathogens among preschool aged children in the MORDOR Niger trial by treatment group and age, 2015-2018. **(A)** Mean IgG responses (MFI-bg). **(B)** Seroprevalence. Shaded area indicates age range included in force of infection analyses. Group means estimated using semiparametric cubic splines. Shaded bands are simultaneous 95% confidence intervals. Created with notebook https://osf.io/smwbn .

**SI Figure 8.**
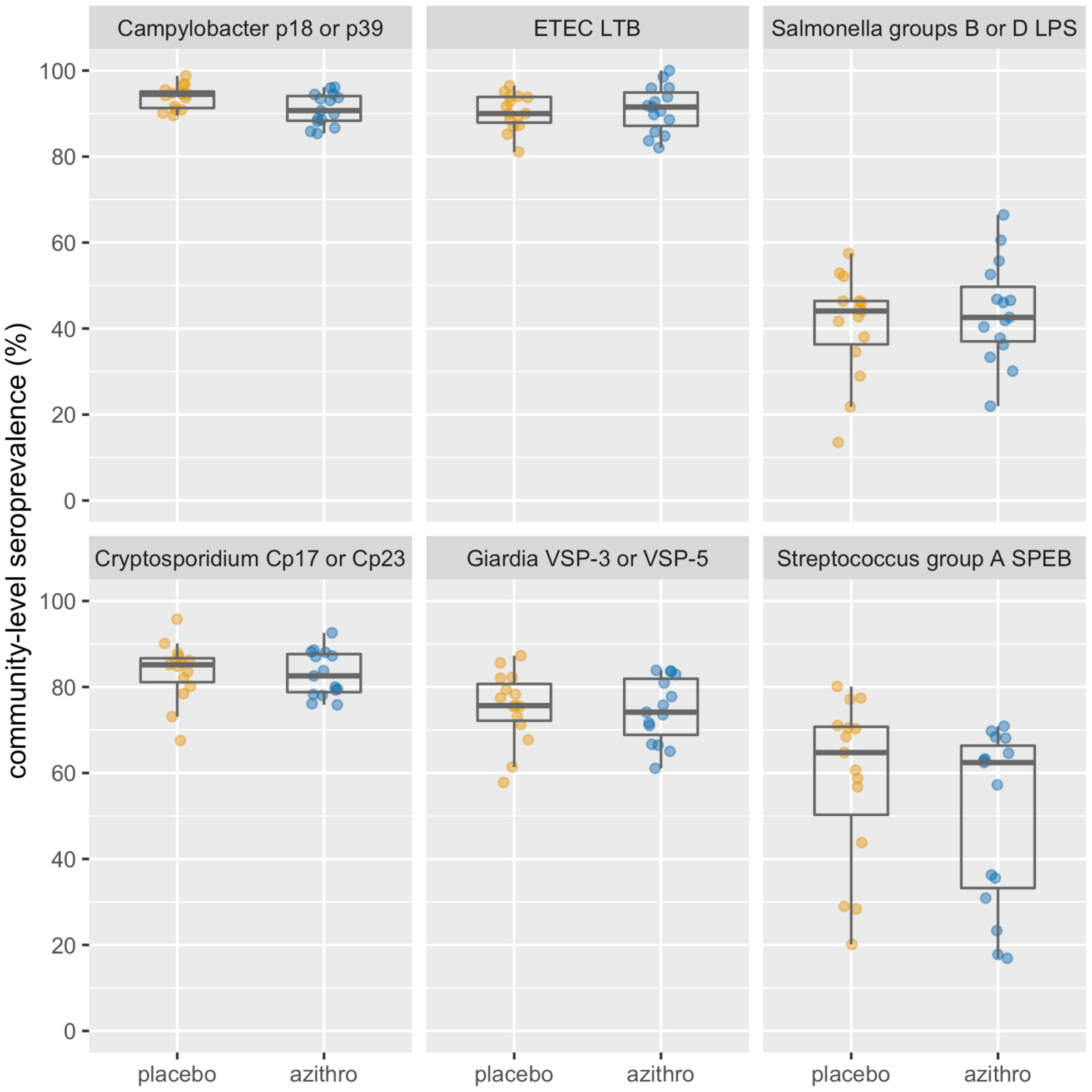
Community level IgG seroprevalence for bacteria and protozoan pathogens among preschool aged children in the MORDOR Niger trial by treatment group, 2015-2018. Box plots show the median and interquartile range for the 15 communities in each group. Created with notebook https://osf.io/b2v3r.

**SI Figure 9.**
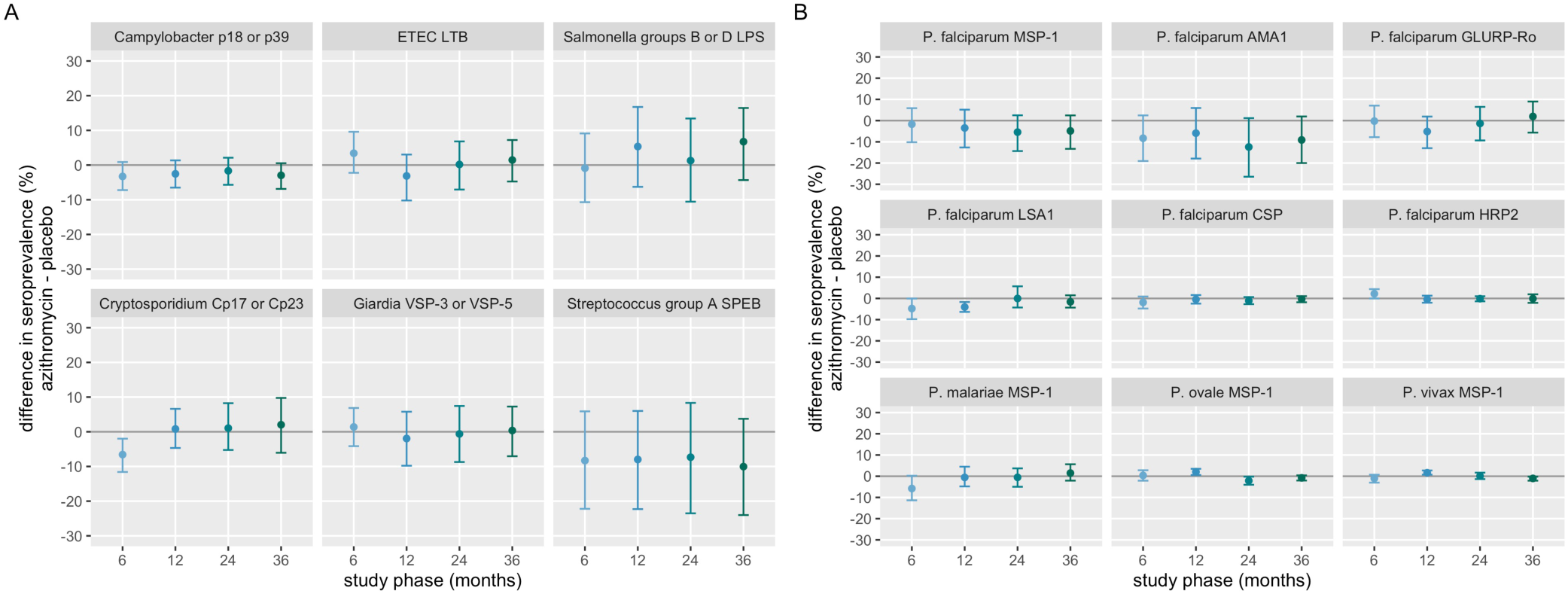
Difference in seroprevalence between intervention groups for bacterial and protozoan pathogens and malaria among children under age five years in MORDOR Niger, 2015-2018, stratified by study phase (months since baseline). (**A**) Bacteria and protozoan antibody responses. (**B**) Malaria antibody responses. There was no evidence for additive effect modification by study phase. Created with notebook https://osf.io/w2rvp, which includes additional details including formal tests of additive effect modification.

**SI Figure 10.**
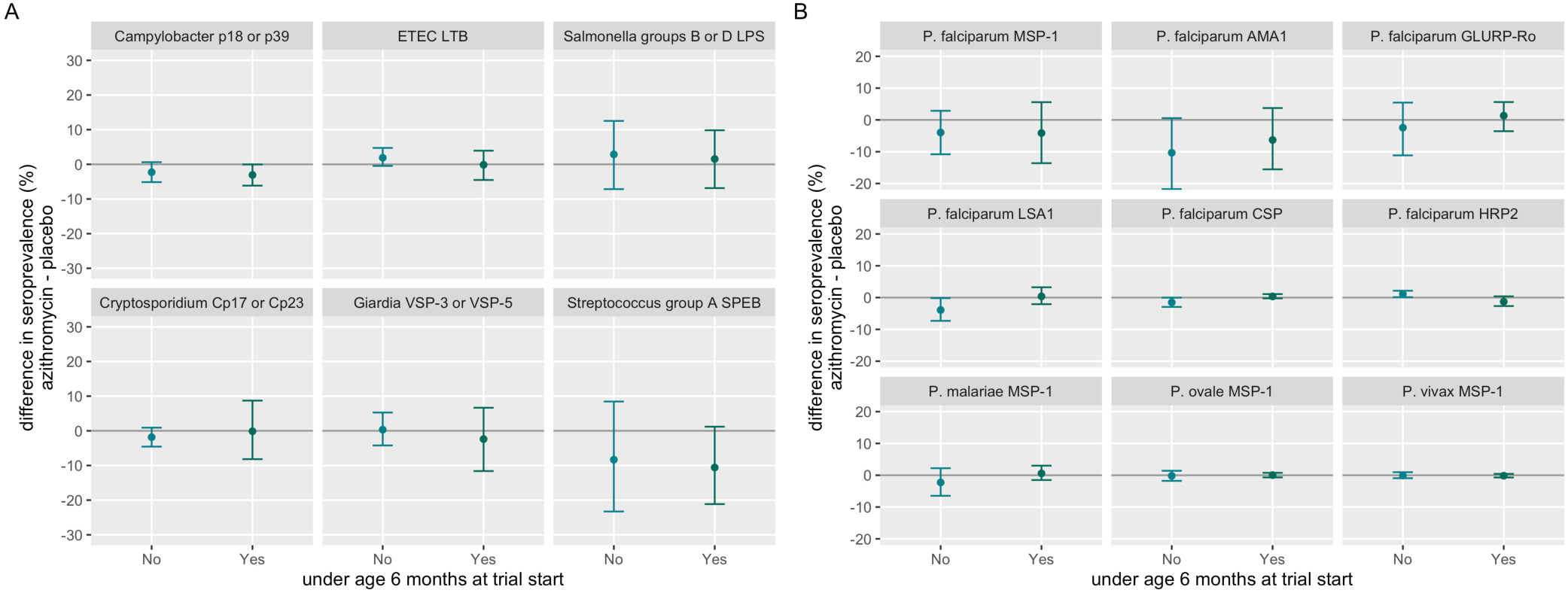
Difference in seroprevalence between intervention groups for bacterial and protozoan pathogens and malaria among children under age five years in MORDOR Niger, 2015-2018, stratified by child age at the start of the trial. (**A**) Bacterial and protozoan antibody responses. (**B**) Malaria antibody responses. There was no evidence for additive effect modification by child age. Created with notebook https://osf.io/w2rvp, which includes additional details including formal tests of additive effect modification.

**SI Table 1.** Seroconversion rates for malaria, bacterial, and protozoan pathogens estimated in longitudinal analyses of children under age five years in MORDOR Niger, 2015-2018. Seroconversion was defined as transition from seronegative to seropositive status. IRR: incidence rate ratio for azithromycin / placebo seroconversion rates. Created with notebook https://osf.io/9875t.

| Pathogen, antigen | Azithromycin |  |  | Placebo |  |  | IRR (95% CI) |
| --- | --- | --- | --- | --- | --- | --- | --- |
|  | N events | Person-Years | Rate per year (95% CI) | N events | Person-Years | Rate per year (95% CI) |  |
| Malaria |  |  |  |  |  |  |  |
| P. falciparum MSP-1 | 88 | 173.58 | 0.51 (0.41, 0.64) | 76 | 107.04 | 0.71 (0.46, 1.02) | 0.71 (0.48, 1.16) |
| P. falciparum AMA1 | 94 | 345.75 | 0.27 (0.19, 0.38) | 106 | 192.96 | 0.55 (0.41, 0.76) | 0.49 (0.31, 0.76) |
| P. falciparum GLURP-Ro | 69 | 606.92 | 0.11 (0.08, 0.15) | 73 | 469.96 | 0.16 (0.11, 0.22) | 0.73 (0.44, 1.13) |
| P. falciparum LSA1 | 18 | 684.29 | 0.03 (0.01, 0.06) | 25 | 567.88 | 0.04 (0.02, 0.07) | 0.60 (0.15, 1.50) |
| P. falciparum CSP | 9 | 722.33 | 0.01 (0.00, 0.02) | 14 | 588.29 | 0.02 (0.01, 0.04) | 0.52 (0.19, 1.14) |
| P. falciparum HRP2 | 13 | 720.21 | 0.02 (0.01, 0.03) | 9 | 596.46 | 0.02 (0.01, 0.02) | 1.20 (0.47, 3.60) |
| P. malariae MSP-1 | 34 | 676.12 | 0.05 (0.02, 0.08) | 30 | 563.88 | 0.05 (0.03, 0.07) | 0.95 (0.43, 1.77) |
| P. ovale MSP-1 | 13 | 723.12 | 0.02 (0.01, 0.03) | 11 | 602.21 | 0.02 (0.01, 0.03) | 0.98 (0.38, 2.43) |
| P. vivax MSP-1 | 4 | 723.46 | 0.01 (0.00, 0.01) | 9 | 604.00 | 0.01 (0.01, 0.02) | 0.37 (0.00, 1.07) |
| Bacteria & protozoa |  |  |  |  |  |  |  |
| Campylobacter p18 or p39 | 63 | 29.21 | 2.16 (1.85, 2.51) | 34 | 12.79 | 2.66 (2.18, 3.25) | 0.81 (0.62, 1.04) |
| ETEC LTB | 69 | 33.71 | 2.05 (1.68, 2.57) | 51 | 22.67 | 2.25 (1.85, 2.59) | 0.91 (0.71, 1.22) |
| Salmonella groups B or D LPS | 162 | 432.21 | 0.37 (0.28, 0.50) | 121 | 403.54 | 0.30 (0.21, 0.39) | 1.25 (0.81, 1.94) |
| Cryptosporidium Cp17 or Cp23 | 80 | 63.42 | 1.26 (0.98, 1.54) | 53 | 45.67 | 1.16 (0.83, 1.47) | 1.09 (0.76, 1.57) |
| Giardia VSP-3 or VSP-5 | 87 | 81.50 | 1.07 (0.81, 1.36) | 53 | 53.04 | 1.00 (0.77, 1.28) | 1.07 (0.74, 1.54) |
| Streptococcus group A SPEB | 140 | 367.58 | 0.38 (0.22, 0.59) | 120 | 288.12 | 0.42 (0.29, 0.61) | 0.91 (0.45, 1.59) |

**SI Table 2.** Seroreversion rates for malaria, bacterial, and protozoan pathogens estimated in longitudinal analyses of preschool children in MORDOR Niger, 2015-2018. Seroreversion was defined as a transition from seropositive to seronegative status. IRR: incidence rate ratio for azithromycin / placebo seroreversion rates. The IRR was not estimated for ETEC LTB because there were no events in the placebo group. Created with notebook https://osf.io/9875t.

| Pathogen, antigen | Azithromycin |  |  | Placebo |  |  | IRR (95% CI) |
| --- | --- | --- | --- | --- | --- | --- | --- |
|  | N events | Person-Years | Rate per year (95% CI) | N events | Person-Years | Rate per year (95% CI) |  |
| Malaria |  |  |  |  |  |  |  |
| P. falciparum MSP-1 | 42 | 493.04 | 0.09 (0.06, 0.11) | 35 | 439.25 | 0.08 (0.05, 0.11) | 1.07 (0.65, 1.80) |
| P. falciparum AMA1 | 35 | 320.71 | 0.11 (0.07, 0.17) | 38 | 336.12 | 0.11 (0.08, 0.16) | 0.97 (0.55, 1.67) |
| P. falciparum GLURP-Ro | 34 | 81.58 | 0.42 (0.29, 0.60) | 28 | 87.71 | 0.32 (0.21, 0.46) | 1.31 (0.77, 2.19) |
| P. falciparum LSA1 | 20 | 38.25 | 0.52 (0.36, 0.91) | 21 | 22.92 | 0.92 (0.67, 1.24) | 0.57 (0.36, 1.05) |
| P. falciparum CSP | 12 | 9.79 | 1.23 (0.77, 1.58) | 14 | 10.92 | 1.28 (0.86, 1.80) | 0.96 (0.51, 1.59) |
| P. falciparum HRP2 | 12 | 9.75 | 1.23 (0.60, 1.85) | 12 | 6.00 | 2.00 (1.29, 3.47) | 0.62 (0.27, 1.15) |
| P. malariae MSP-1 | 25 | 38.58 | 0.65 (0.40, 1.08) | 25 | 21.62 | 1.16 (0.67, 1.66) | 0.56 (0.30, 1.16) |
| P. ovale MSP-1 | 15 | 8.08 | 1.86 (1.13, 2.79) | 2 | 2.58 | 0.77 (0.00, 1.20) | 2.40 (1.21, 4.84) |
| P. vivax MSP-1 | 13 | 9.96 | 1.31 (0.88, 1.87) | 3 | 2.21 | 1.36 (1.20, 1.60) | 0.96 (0.64, 1.40) |
| Bacteria & protozoa |  |  |  |  |  |  |  |
| Campylobacter p18 or p39 | 3 | 103.62 | 0.03 (0.00, 0.06) | 1 | 93.50 | 0.01 (0.00, 0.04) | 2.71 (0.00, 5.33) |
| ETEC LTB | 2 | 96.62 | 0.02 (0.00, 0.05) | 0 | 76.33 | 0 | n/a |
| Salmonella groups B or D LPS | 56 | 298.50 | 0.19 (0.12, 0.28) | 40 | 209.62 | 0.19 (0.14, 0.27) | 0.98 (0.54, 1.66) |
| Cryptosporidium Cp17 or Cp23 | 24 | 55.29 | 0.43 (0.28, 0.63) | 7 | 45.92 | 0.15 (0.05, 0.25) | 2.85 (1.52, 8.49) |
| Giardia VSP-3 or VSP-5 | 11 | 38.92 | 0.28 (0.10, 0.61) | 2 | 44.00 | 0.05 (0.00, 0.13) | 6.22 (1.33, 21.06) |
| Streptococcus group A SPEB | 55 | 388.79 | 0.14 (0.11, 0.21) | 39 | 328.25 | 0.12 (0.06, 0.21) | 1.19 (0.62, 2.61) |

**SI Table 3.** **Homology for *Campylobacter jejuni* p18 and p39 antigens.** Results from a Basic Local Alignment Search Tool for Proteins (BLASTP) analysis.

| Species | p18 (cj0113/ Omp18/ PAL) |  |  |  | p39 (amino acids 188-508 of cj0017c/ dsbl) |  |  |  |
| --- | --- | --- | --- | --- | --- | --- | --- | --- |
|  | Strain | Accession No. | Identity (%) | Similarity (%) | Strain | Accession No. | Identity (%) | Similarity (%) |
| *Campylobacter coli* | PNUSAC008160 | EAH9852560 | 100<sup>+</sup> | 100 | PNUSAC008160 | EAH9851771 | 99 | 99 |
| *Campylobacter lari* | 2003D-6133 | EAI4455352 | 100<sup>+</sup> | 100 | PNUSAC009717 | EBF6097769 | 100 | 100 |
| *Campylobacter upsaliensis* | AG20-0002 | EDP7905829 | 88<sup>+</sup> | 94 | PNUSAC003222 | EAJ2130115 | 80 | 89 |
| *Campylobacter fetus* | PNUSAC002822 | EAK5660503 | 94<sup>+</sup> | 98 | PNUSAC008973 | EAI3655429 | 99 | 99 |
| *Campylobacter hyointestinalis* | PNUSAC003488 | EAK5450665 | 100<sup>+</sup> | 100 | PNUSAC002793 | EAI7421273 | 92 | 95 |
| *Campylobacter helveticus* |  | WP_082199026 | 88<sup>+</sup> | 93 |  | WP_131937627 | 79 | 88 |
| *Campylobacter rectus* |  | WP_124851039 | 56<sup>+</sup> | 72 |  | WP_004319693 | 71 | 83 |
| *Campylobacter mucosalis* |  | WP_169764118 | 55<sup>w</sup> | 77 |  | WP_171993456 | 64 | 77 |
| *Campylobacter sputorum* |  | WP_033916677 | 56<sup>w</sup> | 73 |  | WP_089182023 | 62 | 77 |
| *Helicobacter pylori* |  | WP_140528957 | 33<sup>nr</sup> | 53 |  | WP_126442681 | 32 | 53 |
| *Haemophilus influenzae* |  | WP_061720771 | 36<sup>nr</sup> | 51 |  | Not found |  |  |
| *Escherichia coli* | NCTC13353 | SQB78611 | 36 | 53 |  | Not found |  |  |
| *Salmonella enterica* | serovar Eastbourne | EBS5740044 | 37 | 56 |  | Not found* |  |  |

Table Footnotes:
Results from Burnens, A., Stucki, U., Nicolet, J. & Frey, J. Identification and characterization of an immunogenic outer membrane protein of *Campylobacter jejuni*. *J. Clin. Microbiol.* **33**, 2826–2832 (1995)
<sup>+</sup> Cross-reactive with *C. jejuni* antibody
<sup>w</sup> Weakly cross-reactive with *C. jejuni* antibody
<sup>nr</sup> Non-reactive with *C. jejuni* antibody
\*Amino acids 1-187 of the dsbl protein are present in the genome, but p39 is not

**Text S1.**
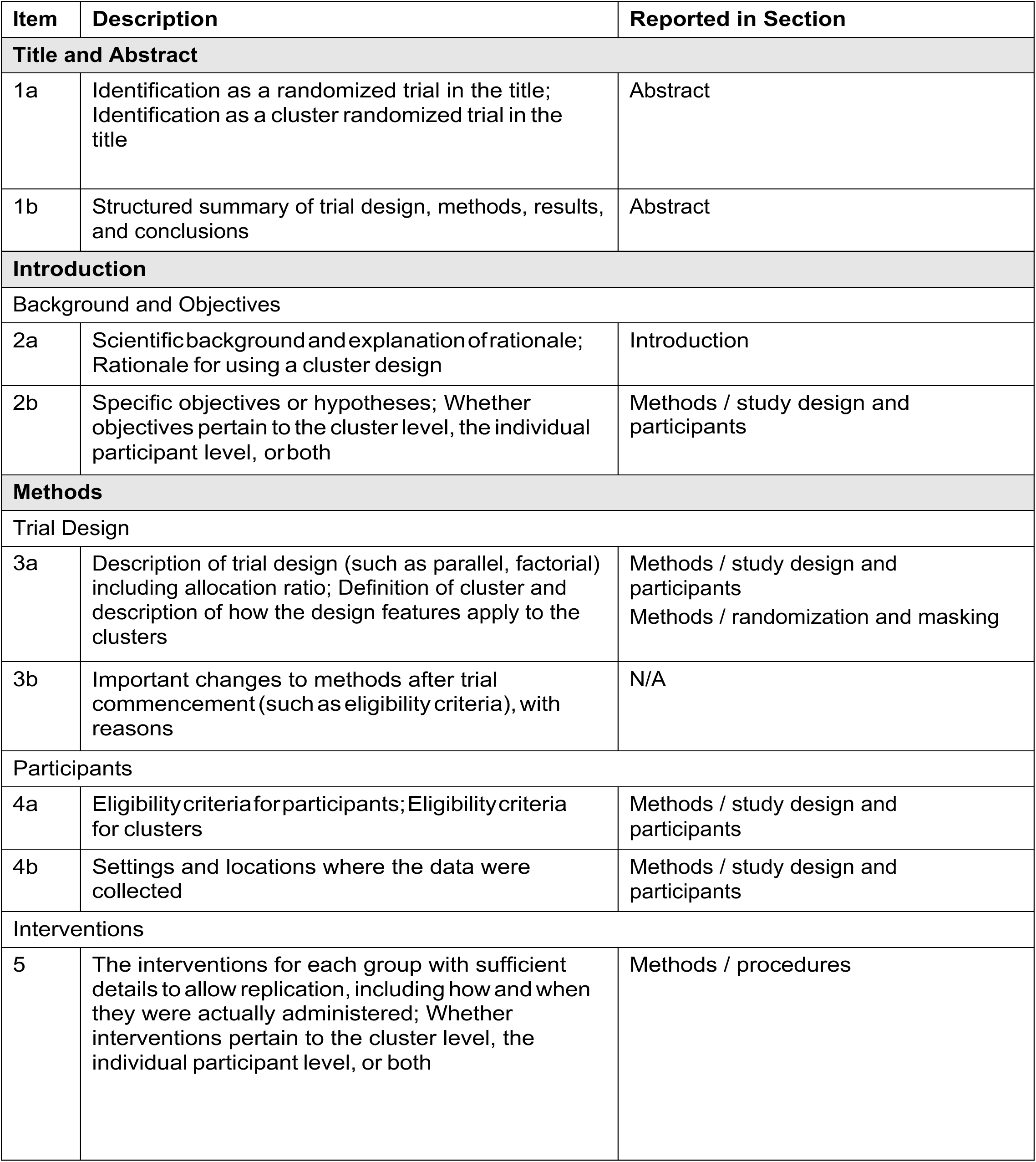
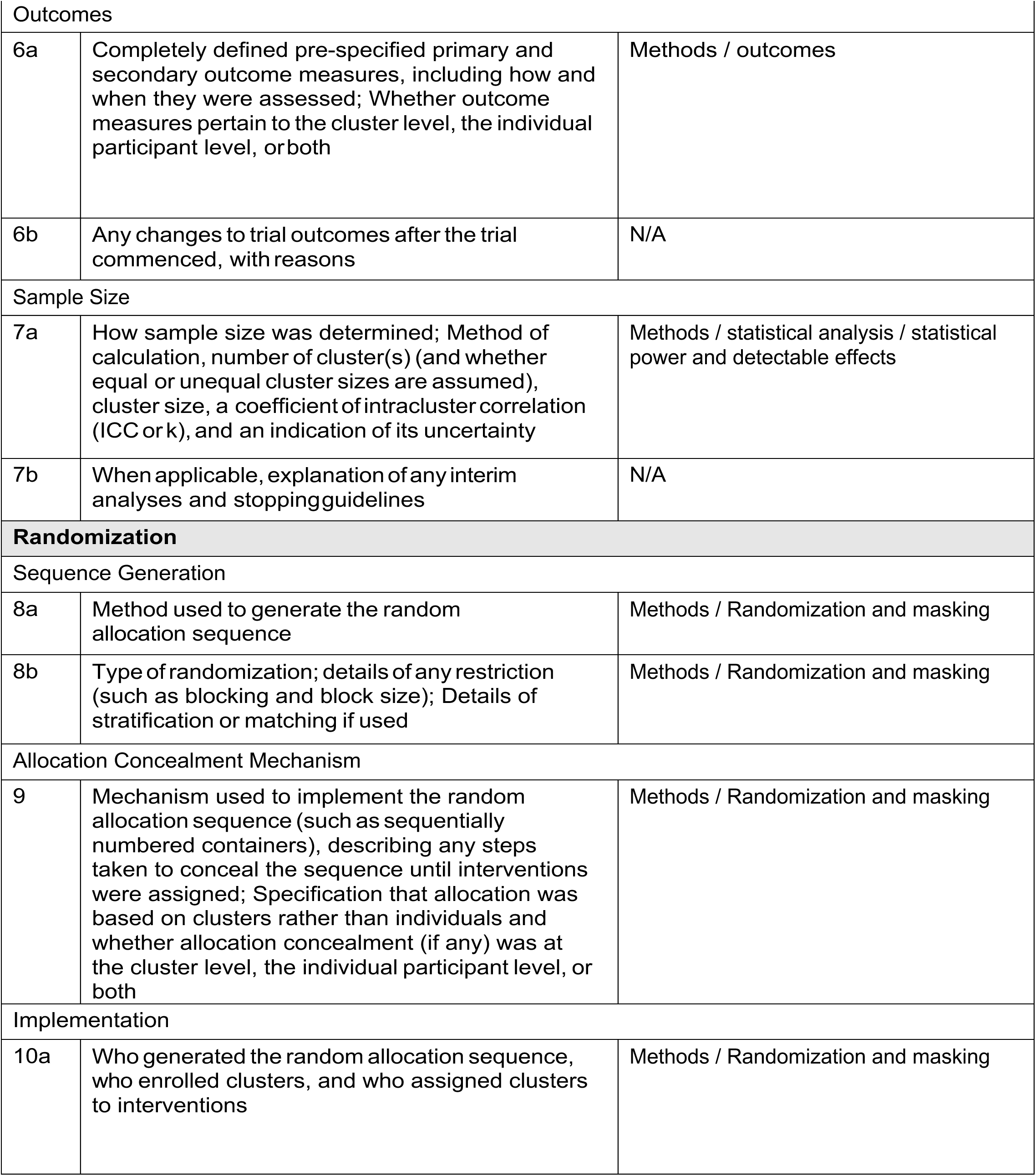
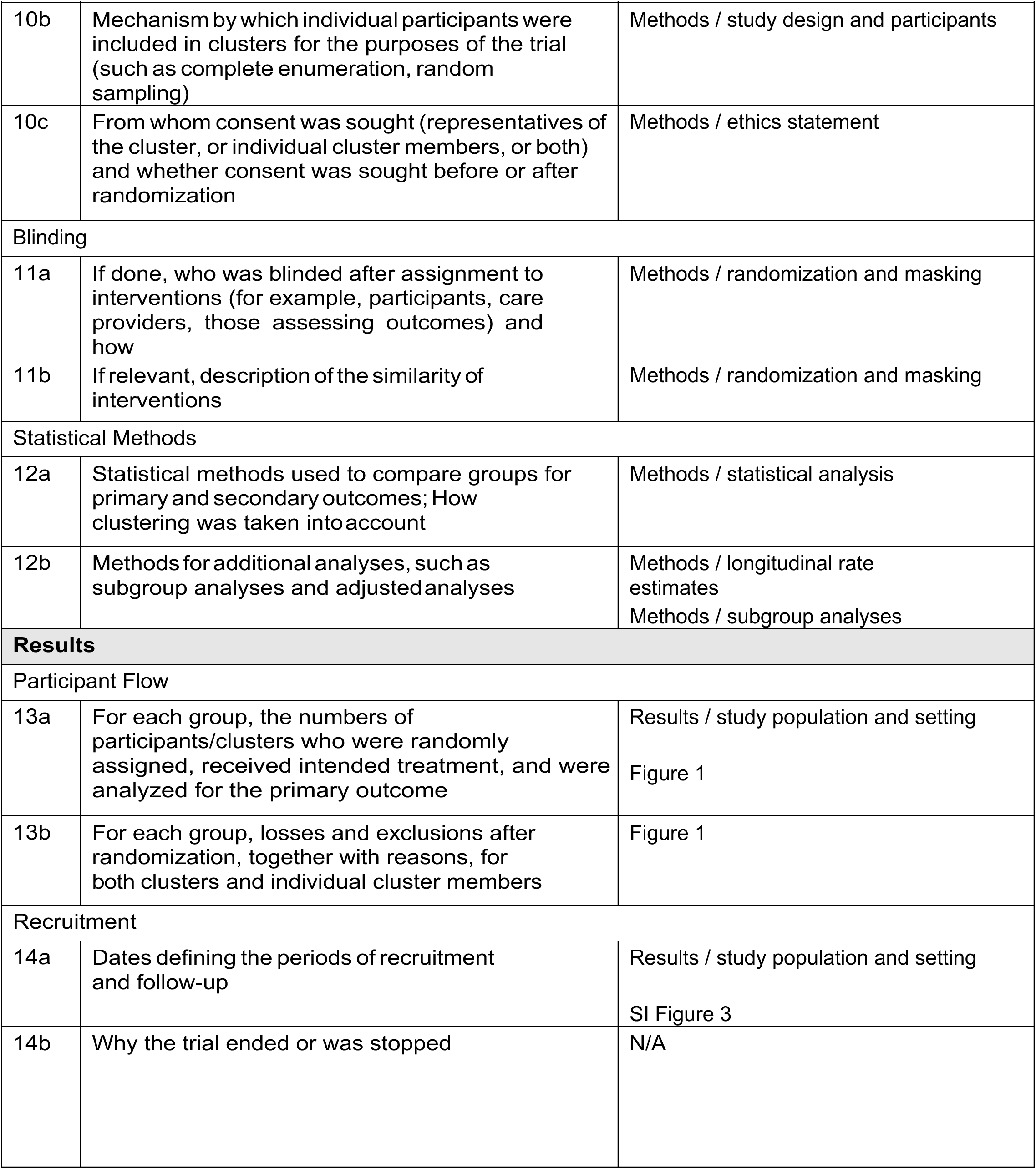
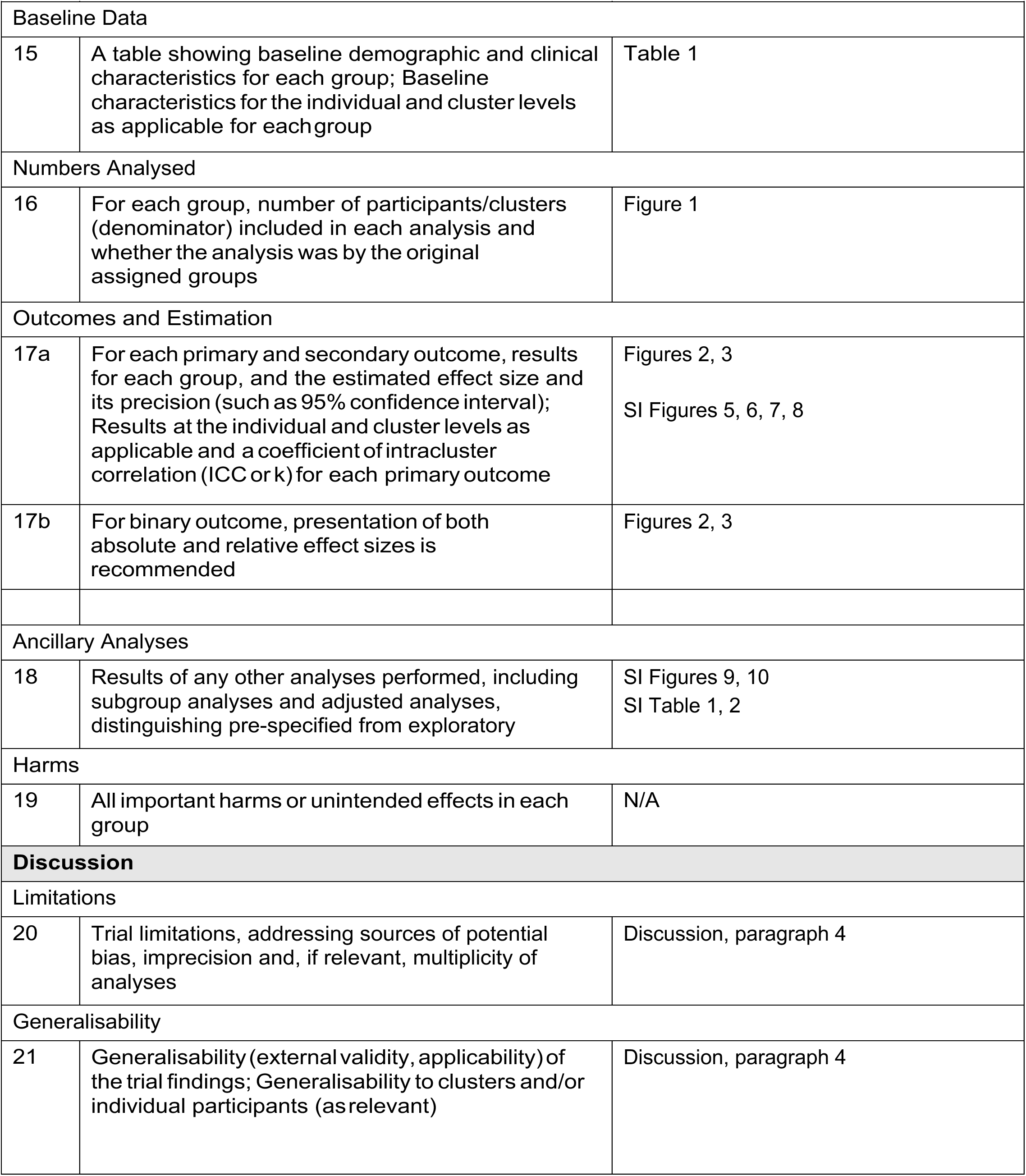
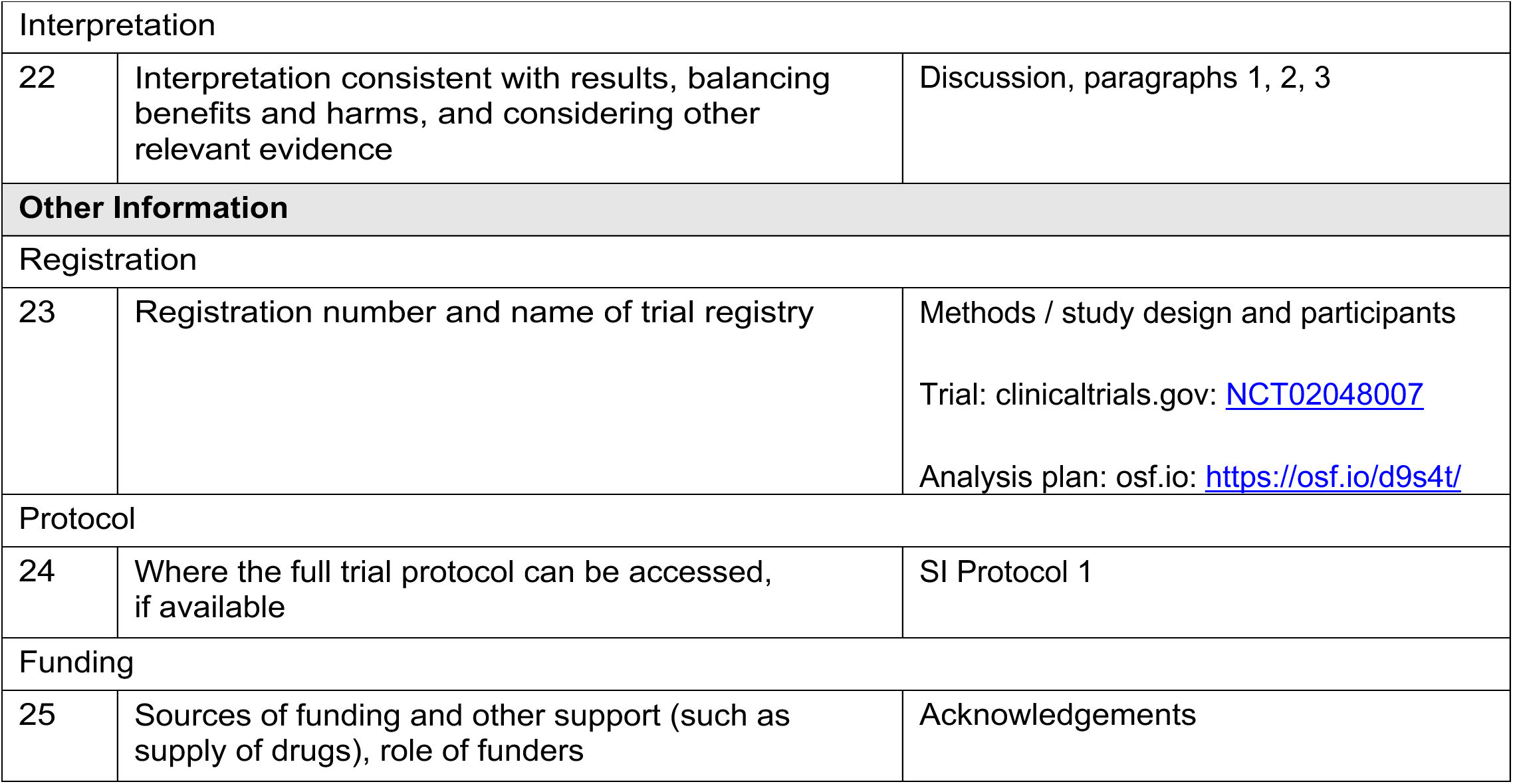
CONSORT checklist.

